# Will Vaccine-derived Protective Immunity Curtail COVID-19 Variants in the US?

**DOI:** 10.1101/2021.06.30.21259782

**Authors:** Marina Mancuso, Steffen E. Eikenberry, Abba B. Gumel

## Abstract

Multiple effective vaccines are currently being deployed to combat the COVID-19 pandemic, and are viewed as the major factor in marked reductions of disease burden in regions with moderate to high vaccination coverage. The effectiveness of COVID-19 vaccination programs is, however, significantly threatened by the emergence of new SARS-COV-2 variants that, in addition to being more transmissible than the wild-type (original) strain, may at least partially evade existing vaccines. A two-strain (one wild-type, one variant) and two-group (vaccinated or otherwise) mechanistic mathematical model is designed and used to assess the impact of the vaccine-induced cross-protective efficacy on the spread the COVID-19 pandemic in the United States. Rigorous analysis of the model shows that, in the absence of any co-circulating SARS-CoV-2 variant, the vaccine-derived herd immunity threshold needed to eliminate the wild-type strain can be achieved if 59% of the US population is fully-vaccinated with either the Pfizer or Moderna vaccine. This threshold increases to 76% if the wild-type strain is co-circulating with the Alpha variant (a SARS-CoV-2 variant that is 56% more transmissible than the wild-type strain). If the wild-type strain is co-circulating with the Delta variant (which is estimated to be 100% more transmissible than the wild-type strain), up to 82% of the US population needs to be vaccinated with either of the aforementioned vaccines to achieve the vaccine-derived herd immunity. Global sensitivity analysis of the model reveal the following four parameters as the most influential in driving the value of the reproduction number of the variant strain (hence, COVID-19 dynamics) in the US: (a) the infectiousness of the co-circulating SARS-CoV-2 variant, (b) the proportion of individuals fully vaccinated (using Pfizer or Moderna vaccine) against the wild-type strain, (c) the cross-protective efficacy the vaccines offer against the variant strain and (d) the modification parameter accounting for the reduced infectiousness of fully-vaccinated individuals experiencing breakthrough infection. Specifically, numerical simulations of the model show that future waves or surges of the COVID-19 pandemic can be prevented in the US if the two vaccines offer moderate level of cross-protection against the variant (at least 67%). This study further suggests that a new SARS-CoV-2 variant can cause a significant disease surge in the US if (i) the vaccine coverage against the wild-type strain is low (roughly < 66%), (ii) the variant is much more transmissible (e.g., 100% more transmissible) than the wild-type strain, or (iii) the level of cross-protection offered by the vaccine is relatively low (e.g., less than 50%). A new SARS-CoV-2 variant will not cause such surge in the US if it is only moderately more transmissible (e.g., the Alpha variant, which is 56% more transmissible) than the wild-type strain, at least 66% of the population of the US is fully vaccinated, and the three vaccines being deployed in the US (Pfizer, Moderna, and Johnson & Johnson) offer a moderate level of cross-protection against the variant.

## 1. Introduction

Since its emergence late in December 2019, the novel coronavirus pandemic (COVID-19), caused by the RNA virus SARS-COV-2, has spread to every corner of the world. While other coronavirus pandemics have emerged in recent decades (including the 2002-2004 severe acute respiratory syndrome (SARS-CoV-1) and the 2012-present middle eastern respiratory syndrome (MERS-CoV) pandemics (60)), COVID-19 represents the greatest global public health challenge since the 1918-1919 influenza pandemic. As of August 3, 2021, there have been over 198 million confirmed COVID-19 cases and over 4.2 million deaths worldwide, with the US being the most affected country (with over 35 million confirmed cases and now over 613,000 COVID-19 deaths)(21).

The primary mode of transmission of COVID-19 is via respiratory droplets (16), and the most common symptoms of the disease include dry cough, shortness of breath, fever, fatigue, muscle aches, and loss of taste or smell. The symptoms typically develop within 2-14 days after contacting the SARS-CoV-2 virus, and can last for up to two weeks (although a small number of infected individuals experience “long COVID-19,” when symptoms endure for several weeks or months, and interfere with day-to-day life (17)). The severity of symptoms varies widely from wholly asymptomatic disease to severe multi-organ system failure and death. Risk of a poor outcome increases with comorbidities, such as immunocompromised status, heart or lung disease, diabetes, etc., while the mortality rate increases exponentially with age. Transmission to susceptible individuals peaks early in the natural history of the disease, with infected individuals infectious several days before the onset of symptoms (pre-symptomatic stage). Infected individuals who never develop symptoms may also still transmit the disease as asymptomatic carriers (18).

Until late December 2020, control and mitigation measures against COVID-19 in the US were exclusively non-pharmaceutical interventions (NPIs), such as the quarantine of those suspected of being exposed to the disease, isolation of individuals with disease symptoms, community “lockdowns,” social (physical)-distancing, and the use of face masks (29; 59; 46; 31; 38; 40). Numerous states and jurisdictions within the US implemented lockdown and mask mandates of varying strictness and duration to limit COVID-19 spread (28; 61; 33; 79; 80; 81; 35; 49; 1; 2).

The US Food and Drug Administration (FDA) gave emergency use authorization (EUA) for the first vaccine against COVID-19 on December 11, 2020 (72; 74). This vaccine was developed by Pfizer-BioNtech and based on delivery of RNA (mRNA) encoding the SARS-CoV-2 spike protein (54). Two doses of the vaccine administered 21 days apart showed 95% efficacy against symptomatic COVID-19 in clinical trials with 44,000 participants (74; 72). A week later (December 18, 2020), the FDA authorized a vaccine developed by Moderna Inc. (71). This vaccine is also based on mRNA technology. Delivered in two doses administered four weeks apart, the vaccine showed efficacy of 94.5% in initial clinical trials (71). The most recent vaccine to receive EUA (February 27, 2021) is the Janssen vaccine, developed by Johnson & Johnson and is administered as a single dose (73). Unlike the mRNA-based Pfizer and Moderna vaccines, the Johnson & Johnson vaccine was developed based on using adenovirus vector encoding the SARS-CoV-2 spike protein. Results from clinical trials showed the Johnson & Johnson vaccine to be 67% effective in preventing moderate to severe COVID-19 14 days post-vaccination (73). Although the vaccines have generally only been available to those 16 and older (Pfizer) or 18 and older (Moderna, J&J), but the FDA recently extended EUA for the Pfizer vaccine to be administered to children of ages 12–15 (75).

Following EUA in December 2020, limited vaccine doses were made available according to prioritization schedules that varied somewhat by state. Vaccination priority was given to the elderly, healthcare workers, and long-term care residents (12). Vaccinations rates had increased linearly for the first few months of administration, but had since peaked in April 2021 and are on the decline as of July 2021 (13). Even with all individuals aged 12 and above now eligible to receive the Pfizer vaccine in the US, increasing vaccine coverage is now widely viewed as a problem of limited demand rather than supply. While vaccines are highly effective against the original SARS-CoV-2 strain, sustainable control of the COVID-19 epidemic may hinge on the characteristics of emerging variant strains.

Viruses are subject to mutations when replicating, and mutations that increase the fitness of the virus tend to persist in the population. In fact, by the middle of March 2020, the dominant SARS-CoV-2 strain in the US had already mutated from the original strain that emerged in Wuhan, China three months earlier (50). Since then, several additional variants from the original SARS-CoV-2 strain have appeared around the globe (15), with many likely more transmissible (and possibly more deadly) than the original SARS-CoV-2 strain (25; 77). For instance, the B.1.1.7 variant (known as the Alpha variant) first appeared in the United Kingdom during late summer of 2020 (77), and may be associated with an increased risk of disease-induced death (15). The mutations of this variant were found on the spike protein of the SARS-CoV-2 virus (19). This variant was first detected in the US on December 29, 2020 in Colorado (22). The B.1.1.7. variant was shown to be 56% more transmissible than the wild-type (original) SARS-CoV-2 strain (25). Individuals infected with the B.1.1.7. variant were 1.4 times more likely to be hospitalized, and 1.58-1.71 times more likely to die from COVID-19 complications (45). Another variant, B.1.351, emerged independently from the B.1.1.7. variant, but shares some similar mutations (15). The B.1.351 variant was first identified in South Africa in October 2020, and was detected in the US at the end of January 2021 (63). Furthermore, the P.1. SARS-CoV-2 variant emerged out of Brazil. This variant, which has 17 unique mutations, also appeared in the US at the end of January 2021 (57). Finally, the most recent variant to have emerged in the US is the B.1.617.2 (or Delta) variant, that first appeared in India (19).

Other SARS-CoV-2 variants have appeared in RNA sequencing, but the Alpha (B.1.1.7) and Delta (B.1.617.2) variants represent the greatest proportion of variants that are currently circulating in the US, and are currently classified as variants of concern (VOC) (20; 19). VOCs have shown evidence of (i) increased disease transmission, (ii) more severe disease, (iii) evading current diagnostic tests, and/or (iv) resistance to naturally-acquired or vaccine-induced immunity, when compared to the wild-type SARS-CoV-2 strain, which does not contain any major mutations (15; 19; 83). These concerns are compounded by the fact that, as more individuals became vaccinated, mask mandates and social-distancing measures were relaxed, or lifted altogether (3; 27; 78). The SARS-CoV-2 strains sequenced in the US during the middle of May 2021 showed the B.1.1.7 variant accounting for nearly 70% of the variants sequenced, 8.1% of the variants were identified as P.1, and 2.5% for the delta variant (20). But, as of July 31, 2021, the Alpha variant was only 2.9% of the sequenced variants, while the Delta variant increased to over 83% (20). The varying proportions of variant transmission, and how they respond to currently available vaccines, add more uncertainty to the effort to effectively suppress or eliminate the pandemic in the US.

Recent studies regarding the cross-protective immunity of existing vaccines on the SARS-CoV-2 VOCs have shown varying results (62; 83; 36; 84). Some studies evaluating the cross-efficacy of the Pfizer-BioNtech vaccine on the B.1.1.7 and B.1.351 variants showed no significant differences of antibody neutralization when two doses of the vaccine were delivered 18-28 days apart (62; 84). However, another study showed that less neutralizing antibodies were produced in individuals who acquired immunity through infection recovery (convalescent immunity) or only received a single dose of the vaccine (62; 4). Another study compared the cross-efficacy of the B.1.1.7 and B.1.351 variants when two doses of the Moderna vaccine was administered 28 days apart (83), which showed reduction in antibody neutralization for the B.1.351 strain, but not the B.1.1.7 strain. Further research is needed to validate and confirm these initial reports.

Despite the uncertainty regarding the level of cross-efficacy that current EUA vaccines provide against VOCs, some researchers showed that there is, at least, some degree of cross-protection (70; 15). However, it is of major public health interest to understand if the current vaccination program will be enough to eliminate the pandemic in the United States, under a plausible range of variant transmissibility and vaccination cross-protection scenarios.

During the earliest days of the COVID-19 pandemic, before extensive experimental or case data was available, mathematical modeling was a useful tool to gauge the severity and potential impact that the disease would have on the community (42; 44; 51; 64; 66; 32; 85; 68; 31; 23; 48; 29; 46; 59; 38; 40). With the increased availability and administration of the Pfizer, Moderna, and Johnson & Johnson vaccines in the US, as well as uncertainties surrounding the recently emerged SARS-CoV-2 variants, mathematical modeling can be a very useful tool to estimate the extent that cross-protective immunity from vaccines protect the community from the emerging COVID-19 variants. A number of recent modeling studies have used deterministic SEIR-type models to assess the potential impact of SARS-CoV-2 variants on the dynamics of COVID-19 in a population. For instance, Gonzalez-Parra et al. (34) used a two-strain SEIR modeling framework, which also includes disease transmission by asymptomatically-infectious individuals, to simulate COVID-19 dynamics in Colombia under varying levels of variant infectiousness (in comparison to the wildtype strain). A two-strain vaccination model was used by Betti et al. (8) to address the crucial question of whether a mutant SARS CoV-2 strain (mutant) could undermine vaccination efforts. Specifically, they estimated the time when a variant strain had overtaken the wild-type SARS CoV-2 strain in Ontario, Canada.

The current study uses a mathematical model to assess the impact of vaccination and vaccine-induced cross-protection against the B.1.1.7 and other SARS-CoV-2 variants circulating in the US as of June 2021. The two-strain and two-group model, which takes the form of a deterministic system of nonlinear differential equations, is formulated in Section 2.1. The model is fitted using COVID-19 mortality data, and a key unknown parameter of the model is estimated from the fitting, in Section 2.2. Basic qualitative features of the model, including the derivation of vaccine-derived herd immunity threshold for the US and parameter sensitivity analysis, are discussed in Section 3. Numerical simulations of the model are also presented in this section.

## 2. Methods

### 2.1. Model Formulation

In this section, a two-strain, two-group mechanistic model for the transmission dynamics of two SARS-CoV-2 strains (the wild-type and variant strains) in the US in the presence of vaccination will be formulated. Although multiple SARS-CoV-2 variants are co-circulating in the US, we consider only one variant strain in the model formulation. This is because almost all of the jurisdictions in the US that are currently experiencing major surges of SARS-CoV-2 have a dominant variant among the current co-circulating variant strains (20). In formulating the model, the total population of individuals in the community at time *t*, denoted by *N*(*t*), is stratified into two groups based on vaccination status, namely the total unvaccinated group, denoted by *N*_*U*_ (*t*) and the total vaccinated group denoted by *N*_*V*_ (*t*), so that *N*(*t*) = *N*_*U*_ (*t*) + *N*_*V*_ (*t*). The total unvaccinated population (*N*_*U*_ (*t*)) is further stratified into the mutually-exclusive compartments of individuals that are unvaccinated susceptible (*S*_*U*_), latent or “exposed” (*E*_*i*_), pre-symptomatically infectious (*P*_*i*_), symptomatically infectious (*I*_*i*_), asymptomatically infectious (*A*_*i*_), hospitalized (*H*_*i*_), and recovered (*R*_*i*_), with *i* = 1, 2 (represents the SARS-CoV-2 strain), so that

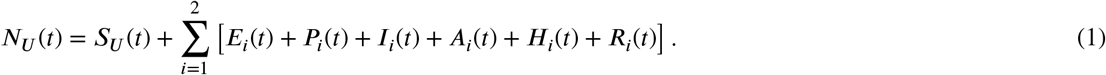

Similarly, the total vaccinated population (*N*_*V*_ (*t*)) is sub-divided into the sub-populations of individuals that are vaccinated susceptible (*S*_*V*_ (*t*)), exposed (*E*_*Vi*_), pre-symptomatically infectious (*P*_*Vi*_), symptomatically infectious (*I*_*Vi*_), asymptomatically infectious (*A*_*Vi*_), hospitalized (*H*_*Vi*_), and recovered (*R*_*Vi*_), with = 1, 2. Thus,

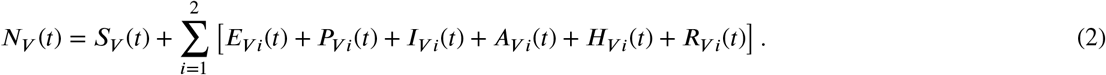

Vaccinated individuals are assumed to have received the full required doses (i.e., they are fully vaccinated). The equations for the rate of change of the aforementioned state variables of the two-strain, two-group vaccination model for COVID-19 transmission dynamics and control in the US are given by the deterministic system of nonlinear differential equations given by Equation (A-8) of Appendix A. The flow diagram of the model is depicted in Figure 1, and the state variables and parameters of the model are described in Tables A.7 and A.8, respectively.

**Figure 1:**
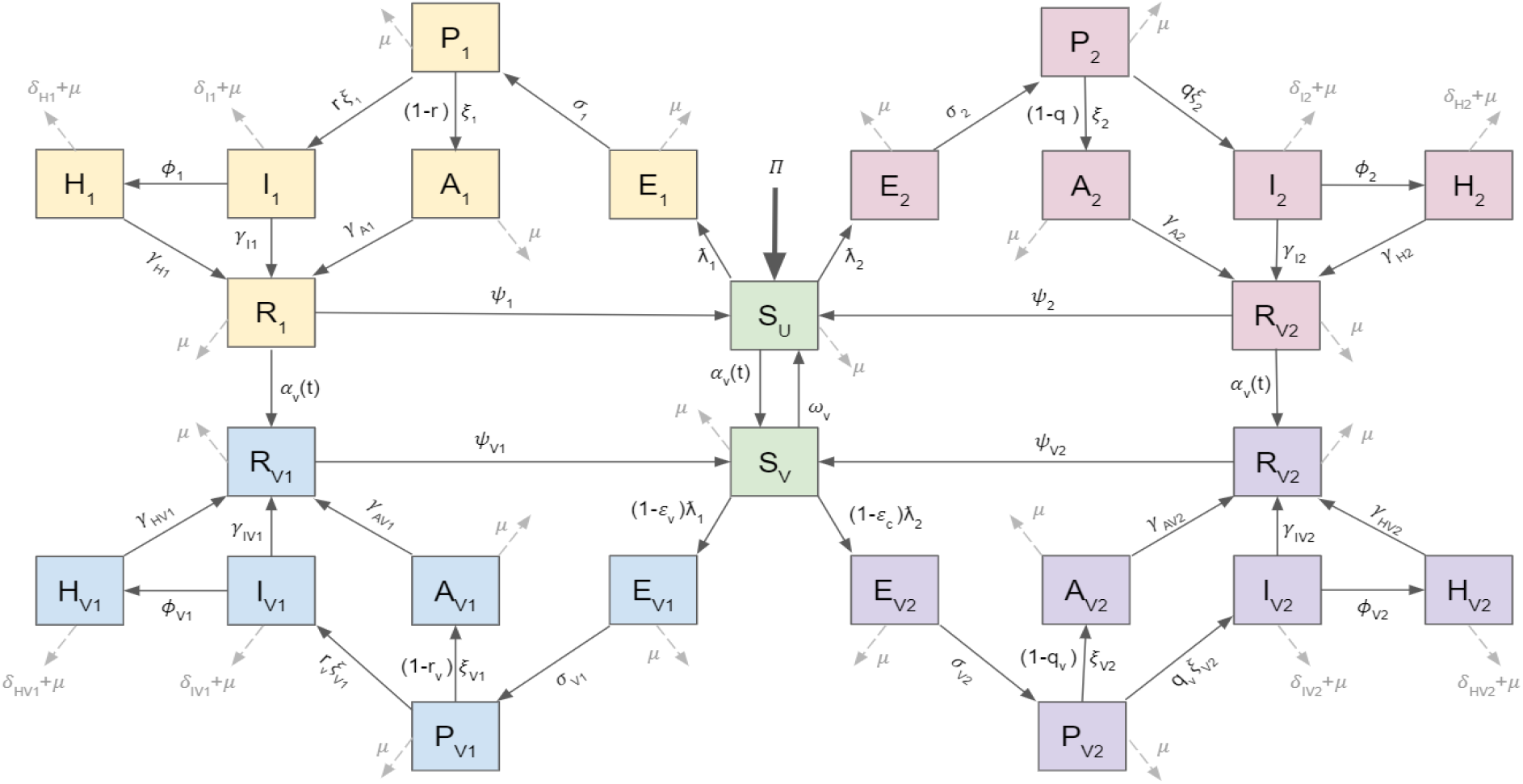
Schematic diagram of the model (A-8).

In the formulation of the model (A-8), we assume a setting where two strains of SARS-CoV-2 are co-circulating in the population, namely:

a. strain 1: this is the wild-type (original) strain. It is assumed to be the predominant strain circulating the US throughout 2020 (50);
b. strain 2: this is a new dominant, variant strain assumed to be circulating in the US since the end of 2020 (22)).

For mathematical tractability and convenience, we assume that susceptible individuals can be infected with either of the two strains, but not with both. Furthermore, we are not aware of real evidence for co-infection of multiple SARS-CoV-2 strains at the current moment. The three EUA vaccines in the US (i.e., the Pfizer, Moderna, and Johnson & Johnson vaccines) target the wild-type strain with efficacy *ε*_*v*_, and are assumed to induce cross-protective efficacy (*ε*_*c*_) against the dominant variant (strain 2) (62; 84; 36; 83). Vaccination is offered to all eligible unvaccinated individuals, but for simplicity, the model does not consider vaccination of currently infected individuals (symptomatic or asymptomatic). In other words, the model only considers vaccinating those who are wholly-susceptible and those who recovered from the disease prior to being vaccinated. Although currently infectious individuals may be getting vaccinated, they comprise of extremely small fraction of the overall population at any given time, and are not included (for simplicity).

Some of the other main assumptions made in the formulation of the model (A-8) include:

i. Homogeneous mixing: the population is well-mixed, such that every member of the community is equally likely to mix with every other member of the community. Further, the waiting times in each epidemiological compartment are exponentially distributed (43).
ii. Because of the inherent age structure in the COVID-19 vaccine administration (where, in the context of the Pfizer vaccine, for instance, only people at age 12 or older are vaccinated (72)), we include demographic (birth and death) parameters to account for the inflow of new susceptible individuals that are eligible for vaccination.
iii. Vaccination is only offered to wholly-susceptible individuals or those who naturally recovered from COVID-19 infection (but not for currently-infected individuals).
iv. Vaccinated individuals are assumed to have received the full recommended dosage (two doses for Pfizer or Moderna vaccine, one dose for the Johnson & Johnson vaccine), and that the vaccine administered was stored at the appropriate temperatures, and that enough time has elapsed for the body to develop immunity.
v. The vaccines are imperfect against infection (i.e, breakthrough infection can occur) (72; 71; 73). The vaccines offer strong therapeutic benefits, *vis a vis* reducing severe disease, hospitalization, and death (72; 71; 73).
vi. Vaccine-derived and natural immunity may wane over time in individuals, in which they revert to the wholly-susceptible class.

### 2.2. Data Fitting and Parameter Estimation

In this section, the model (A-8) is fitted using the cumulative mortality data from Johns Hopkins’ Center for Systems Science & Engineering (from January 22, 2020 to March 6, 2021) (21). Fitting is used to estimate the contact rate parameter for the wild-type strain (strain 1), *β*_1_, during each wave of the COVID-19 pandemic in the US (21; 10). The fitting is done in the absence of vaccination for the period from January 22, 2020 to January 21, 2021, and including vaccination for January 22, 2021 to March 6, 2021. We use a MATLAB routine to minimize the sum of squared differences between each observed cumulative mortality data point and the corresponding mortality point obtained from the model (A-8). COVID-19 mortality data, rather than case data, is used to fit the model since it is more reliable (the latter underestimates the true number of cases owing to the inability to track asymptomatic and pre-symptomatic cases, resulting from the absence of random wide-scale testing across the US) (18). Daily vaccination data, for the period December 21, 2020 to March 6, 2021 (10), was used in the fitting of the model to account for the transition of individuals from nonvaccinated compartments to attaining fully-vaccinated status.

The first case of the Alpha (B.1.1.7) variant was documented in the US on December 29, 2020 (22), although the variant was likely introduced to the US prior to the detection. For the actual fitting of the model, we introduce 1, 000 cases of a variant strain (i.e.,*I*_2_(0) = 1, 000) on December 29, 2020 to estimate variant infections present prior to confirmation of its US detection. Furthermore, vaccination is introduced in the model (A-8) on January 22, 2021. Since our model does not explicitly consider the two-dose vaccine structure or account for any delay from inoculation to (partial) immunity, we introduce a delay from actual to modeled vaccination by constructing an inferred second dose time-series. That is, the available data only gives total doses delivered and does not disaggregate between first and second doses. Therefore, we assume that during the first 30 days of vaccine reporting, all administered doses are first doses. After 30 days, all first doses result in a second dose, and any remaining doses for that day are considered first doses. An inferred second dose time series can thus be constructed iteratively. Second doses were roughly 35% of the total doses administered each day following the first four weeks of vaccination (10).

Furthermore, the following expression was used to determine the time-dependent vaccination rate, *α*_*v*_(*t*):

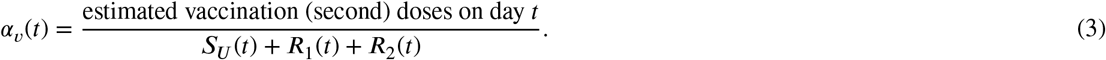

This expression allows for the realistic possibility that individuals who have recovered from infection (and may have naturally-acquired immunity against future infection) will be vaccinated along with susceptible individuals. Thus, a proportion of the vaccination doses administered each day are delivered to wholly-susceptible individuals who have not developed prior COVID-19 immunity.

The result of the data fitting of the model (A-8), using the fixed parameters in Table A.9, is depicted in Figure 2. The left panel of this figure shows the model (A-8) fit using the observed cumulative US mortality data. The right panel shows the simulations of the model (A-8) with the fixed and fitted parameters, compared to the observed daily mortality data for the US. The purple vertical dotted line in each panel indicates the time that a variant strain was introduced into the US population (December 29, 2020). The contact rate for the variant strain (*β*_2_) was assumed to be 56% greater than the contact rate for the wild-type strain (*β*_1_), since the dominant variant circulating the US at the time was the Alpha (B.1.1.7) variant (which is estimated to be at least 56% more transmissible than the original wild-type strain (25)). The brown dotted lines indicate the time when vaccination was introduced into the population (January 22, 2021). The efficacies of the Pfizer and Moderna vaccines against the wild-type strain are set at 94%. That is, *ε*_*v*_ = 0.94 (72; 71). Furthermore, we assumed a moderate level of cross-efficacy of the vaccine against the variant strain, set at 50% (i.e., *ε*_*c*_ = 0.5). The model (A-8), with the chosen fixed parameter values, fits well with the observed cumulative and daily US COVID-19 death data (as shown in Figure 2).

**Figure 2:**
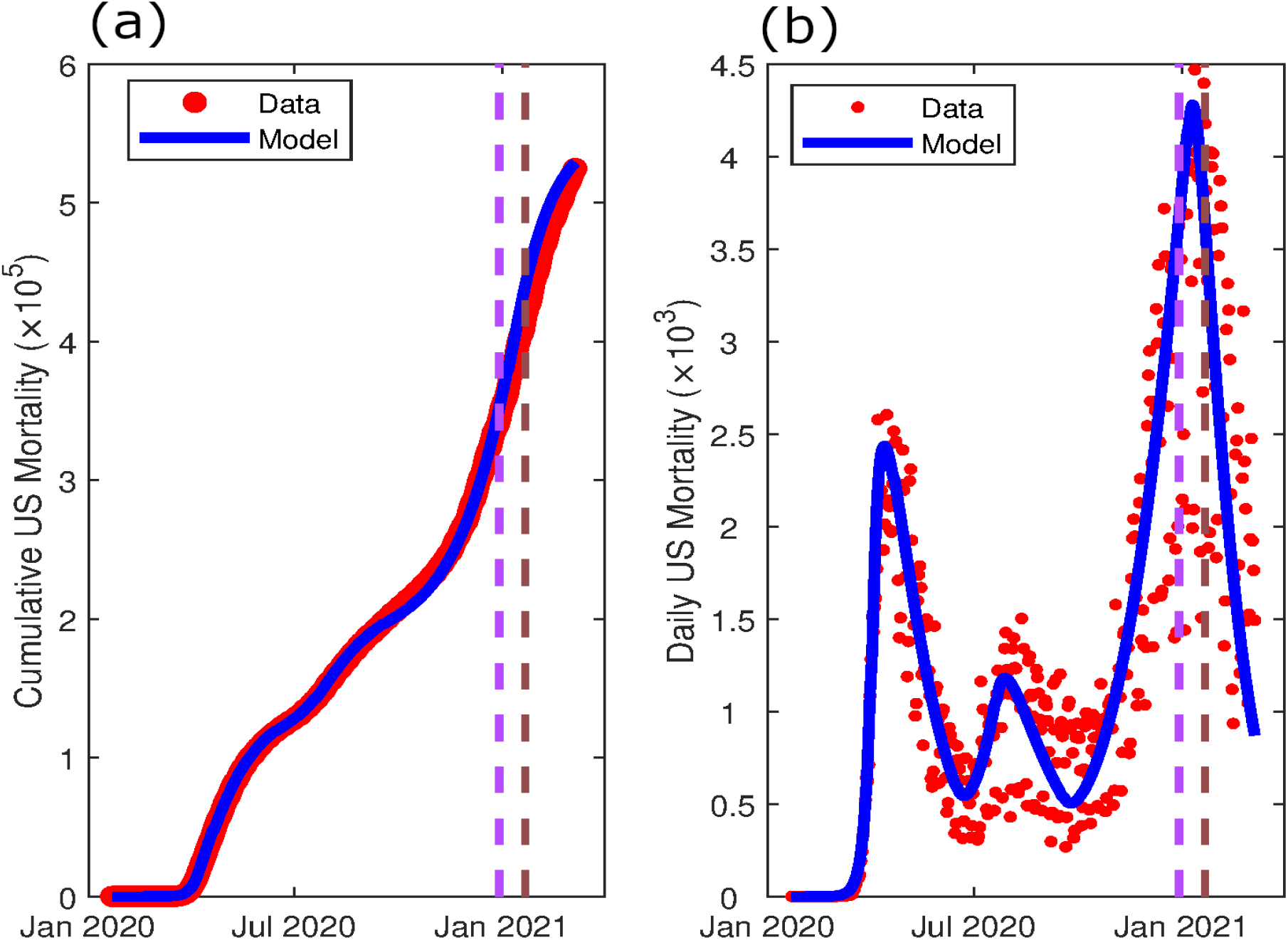
(a) Fitting of the model (A-8) using cumulative mortality data for the US (21). (b) Simulations of the model (A-8) using the fixed and estimated parameters, given in A.9 and strain 1 contact rate values from Table 1, illustrating the daily mortality generated from the model, in comparison to the observed daily mortality data for the US. For both panel, initial conditions used were *S*_*U*_ (0) = 336 million, *I*_1_(0) = 10 and all other state variables set at zero. The purple vertical line shows the time that 1,000 cases of the variant strain (strain 2) was introduced in the population (December 29, 2020), and assumed to be 1.56 times as infectious as strain 1 (25). The brown vertical line shows the time at which vaccination was introduced into the model (January 22, 2021) using data from (10).

The contact rate for the wild-type strain, *β*_1_, varies throughout the course of the pandemic due to implementation and compliance of non-pharmaceutical interventions (NPIs) and vaccination against it. Table 1 shows the values of the contact rate for the wild-type strain (strain 1), *β*_1_, for each period of the pandemic. The first wave of the COVID-19 pandemic (January 22, 2020 to April 1, 2020) was the period with the highest contact rate, and occurred prior to most wide-scale NPI implementation (particularly the CDC’s recommendation of face mask usage). The lockdown period (April 2, 2020 to June 15, 2020) was the period when numerous US states implemented stay-at-home policies, and was shown to have a lower contact rate. The second wave, which occurred during the early to mid summer of 2020 (June 16, 2020 to July 20, 2020) and was period when many states stay-at-home orders expired and businesses started returning to operation. This resulted in a higher contact rate, which in turn lead to more observed COVID-19 mortality. Many states began tightening restrictions on businesses again after seeing the rise in cases and deaths, which resulted in the post-second wave period (July 21, 2020 to September 18, 2020) having a decreased contact rate. As states lifted business restrictions and other COVID-19 related NPIs in mid-September, 2020 (78), and many Americans traveled for the holidays, a large third wave of the pandemic was observed for September 19, 2020 to January 6, 2021, that had a similar contact rate as the second wave. As vaccination distribution commenced towards the final weeks of 2020, the contact rate once again decreased to a level similar to the post-second wave contact rate. We note that during the second and third waves, most US states still maintained a baseline level of NPI implementation and compliance (e.g., social distancing polices, face mask mandates). Thus, the contact rates for the second and third waves were not as high as the initial pandemic wave.

**Table 1.** Results from fitting the contact rate, *β*_1_, to cumulative COVID-19 death data for the US with model (A-8) for the COVID-19 pandemic from January 22, 2020 to March 6, 2021.

| Pandemic Period | $\beta_1$ (per day) | Time Frame |
| --- | --- | --- |
| First wave | 0.3742 | Jan 22, 2020 - April 1, 2020 |
| Lockdown period | 0.0726 | April 2, 2020 - June 15, 2020 |
| Second wave | 0.1494 | June 16, 2020 - July 20, 2020 |
| Post-second wave | 0.0864 | July 21, 2020 - Sept 18, 2020 |
| Third wave | 0.1421 | Sept 19, 2020 - Jan 6, 2021 |
| Post-third wave | 0.0805 | Jan 6, 2021 - March 6, 2021 |

## 3. Results

### 3.1. Mathematical Analysis: Computation of Reproduction Number

The model (A-8) has a unique disease-free equilibrium (DFE) given by

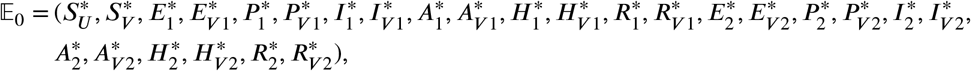

where 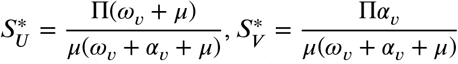 and all other components of 𝔼_0_ are zero. The asymptotic stability property of the disease-free equilibrium will be explored, for the special case of the model where *α*_*v*_(*t*) is constant (i.e., we are considering the *autonomous* version of the model), using the *next generation operator method* (76; 26). Using the notation in (76), it follows that the non-negative matrix (*F*) of new infection terms, and the matrix (*M*) of new transition terms, are given, respectively, by:

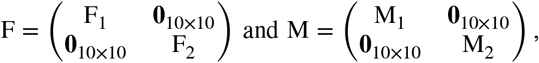

where the matrices F_1_, F_2_, M_1_ and M_2_ are given in Appendix B, and **0**_10×10_ denotes the zero matrix of order 10. It follows that the *vaccination reproduction number* of the autonomous version of the model (A-8), denoted by *ℛ*_*vac*_, is given by (where *ρ* denotes the spectral radius of the next generation matrix FM^−1^):

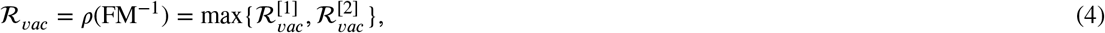

where 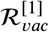 and 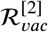 (given in Appendix B) represent, respectively, the constituent reproduction numbers associated with the transmission of strains 1 and 2. The result below follows from Theorem 2 of (76).

#### Theorem 3.1.

*The DFE (*𝔼 _0_*) of the autonomous version of the model (A-8) is locally-asymptotically stable if ℛ*_*vac*_ < 1, *and unstable if ℛ*_*Vac*_ *>* 1.

The threshold quantity, *ℛ*_*vac*_, measures the average number of new COVID-19 cases generated by a single infectious individual introduced into a population where a certain proportion is vaccinated. The epidemiological implication of Theorem 3.1 is that a small influx if COVID-19 cases will not generate an outbreak in the community if the *vaccination reproduction number* (*ℛ*_*vac*_) is brought to (and maintained at) a value less than unity.

### 3.2. Computation of the Vaccine-derived Herd Immunity Threshold

Herd immunity, which is a measure the fraction of susceptible individuals that need to be protected against infection in order to eliminate community transmission of an infectious disease, can be attained through two main ways: natural recovery from infection or from vaccination. However, the safest and fastest way to achieve herd immunity is through vaccination (7; 6). Furthermore, at least 15% of the U.S. population currently cannot be vaccinated due to age restrictions (69). Additionally, individuals who are pregnant, breastfeeding, have underlying medical conditions, as well as for other reasons, may be unable or unwilling to be vaccinated against COVID-19. Thus, it is critical to know what minimum proportion of the susceptible US population need to be vaccinated to achieve vaccine-derived herd immunity (so that the pandemic can be effectively controlled). In this section, we derive the expression for achieving vaccine-derived herd immunity against COVID-19 in the US based on widespread vaccination against the wild-type SARS-CoV-2 strain (i.e., strain 1).

In the absence of vaccination and other public health interventions (e.g., social-distancing, mask wearing, etc), the *vaccination reproduction number* (*ℛ*_*vac*_) reduces to the *basic reproduction number* (denoted by *ℛ*_0_), given below:

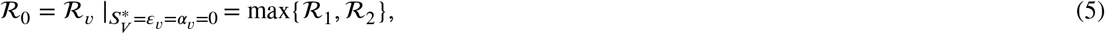

where *ℛ*_1_ and *ℛ*_1_ are the constituent *basic reproduction numbers* for the wild-type (strain 1) and the variant (strain 2) SARS-CoV-2 strains, respectively, given by (where *k*_*i*_, with = 1, 2, ., 20 are given in Appendix B):

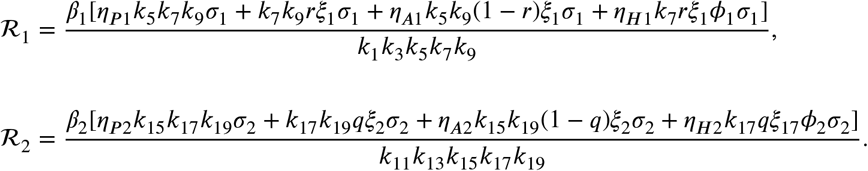

For the case when all individuals in the population are vaccinated, the constituent reproduction numbers for the wildtype (strain 1) and variant (strain 2) SARS-CoV-2 strains become:

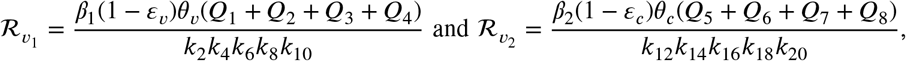

where expressions for *Q*_*i*_ (*i* = 1, 2, ., 8) are also given in Appendix B. Hence, for the case where all individuals in the population are vaccinated, the expression for the vaccination reproduction number of the model is given by:

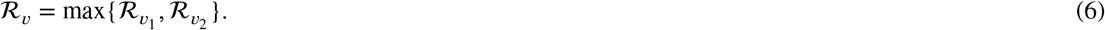

It follows that the *vaccination reproduction numbers* of wild-type (strain 1, 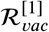) and variant (strain 2, 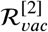) can be re-written in terms of their *basic reproduction numbers ℛ*_1_ and *ℛ*_2_, and reproduction numbers when the entire population is vaccinated, as below:

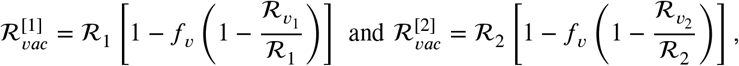

where 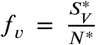 is the proportion of susceptible individuals in the community who have been **fully** vaccinated (at steady-state). The vaccine-derived herd immunity threshold for the wild-type and variant strains are computed by setting 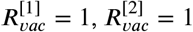 and solving for *f*_*v*_, respectively. We note that it is necessary to ensure, in this computation, that 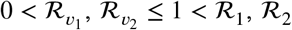, (so that 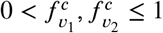). Doing so gives:

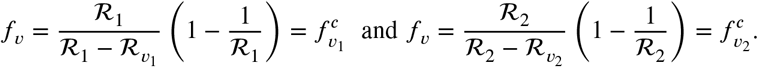

It follows that the vaccine-derived herd immunity threshold for the COVID-19 pandemic in the US, in the presence of both the wild and variant SARS-CoV-2 strains, denoted by 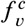, is given by:

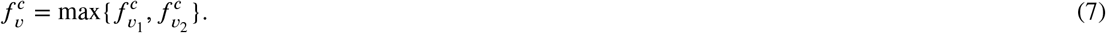

Using the fixed parameters in Table *A*.9 and a contact rate of *β*_1_ = 0.24 per day to account for transmission in the absence of nonpharmaceutical control measures (face mask usage, social distancing policies), it follows from Equation (7) that the vaccine-derived herd immunity threshold for the wild-type (strain 1) SARS-CoV-2 strain (in the absence of the variant strain) is 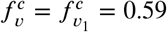 if either the Pfizer or the Moderna vaccine (with efficacy *ε*_*v*_ = 0.94) is used. In other words, herd immunity against the wild-type strain can be achieved in the US, using either the Pfizer or Moderna vaccine, if at least 59% of the US population is fully-vaccinated with any of this vaccine. In this scenario, the herd immunity threshold rises to 64% if only the Johnson & Johnson vaccine (with *ε*_*v*_ = 0.67) is used. Thus, the wild-type SARS-CoV-2 strain can be effectively controlled in the US (in the absence of a co-circulating variant SARS-CoV-2 strain), using any of the three vaccines currently being administered in the US, if 59% (for Pfizer/Moderna vaccine) or 64% (for Johnson & Johnson vaccine) of the US populace is fully vaccinated.

For the case where the wild-type strain is co-circulating with the a dominant SARS-CoV-2 variant that is 56% more transmissible than the wild-type strain (such as the Alpha variant (25)), our study shows that the vaccine-derived herd immunity threshold needed to effectively control both strains is 76% (i.e., 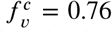) using either the Pfizer or Moderna vaccine, or 93% when using only the Johnson & Johnson vaccine. On the other hand, if the wild-type strain is co-circulating with the Delta variant (which is 100% more transmissible than the wild-type strain), the herd immunity threshold needed to combat both strains increases to 82% if the Pfizer/Moderna vaccine is used.

Figure 3 depicts contour plots of the vaccination reproduction number for the three scenarios with the wild-type strain only (Figure 3 (a)) or in combination with a variant that is 56% (Figure 3 (b)) or 100% ((Figure 3 (c)) more transmissible than the wild-type strain, as a function of vaccine coverage and efficacy. This figure shows that the vaccination reproduction number decreases with increasing values of vaccine coverage and efficacy. This figure further shows that vaccine-derived herd immunity cannot be achieved using the Johnson & Johnson vaccine for the case where the wild-type strain is co-circulating with the Delta variant (Figure 3 (c); since, for the 67% efficacy of this vaccine, the associated reproduction number cannot be brought to a value less than one, even with 100% vaccine coverage).

**Figure 3:**
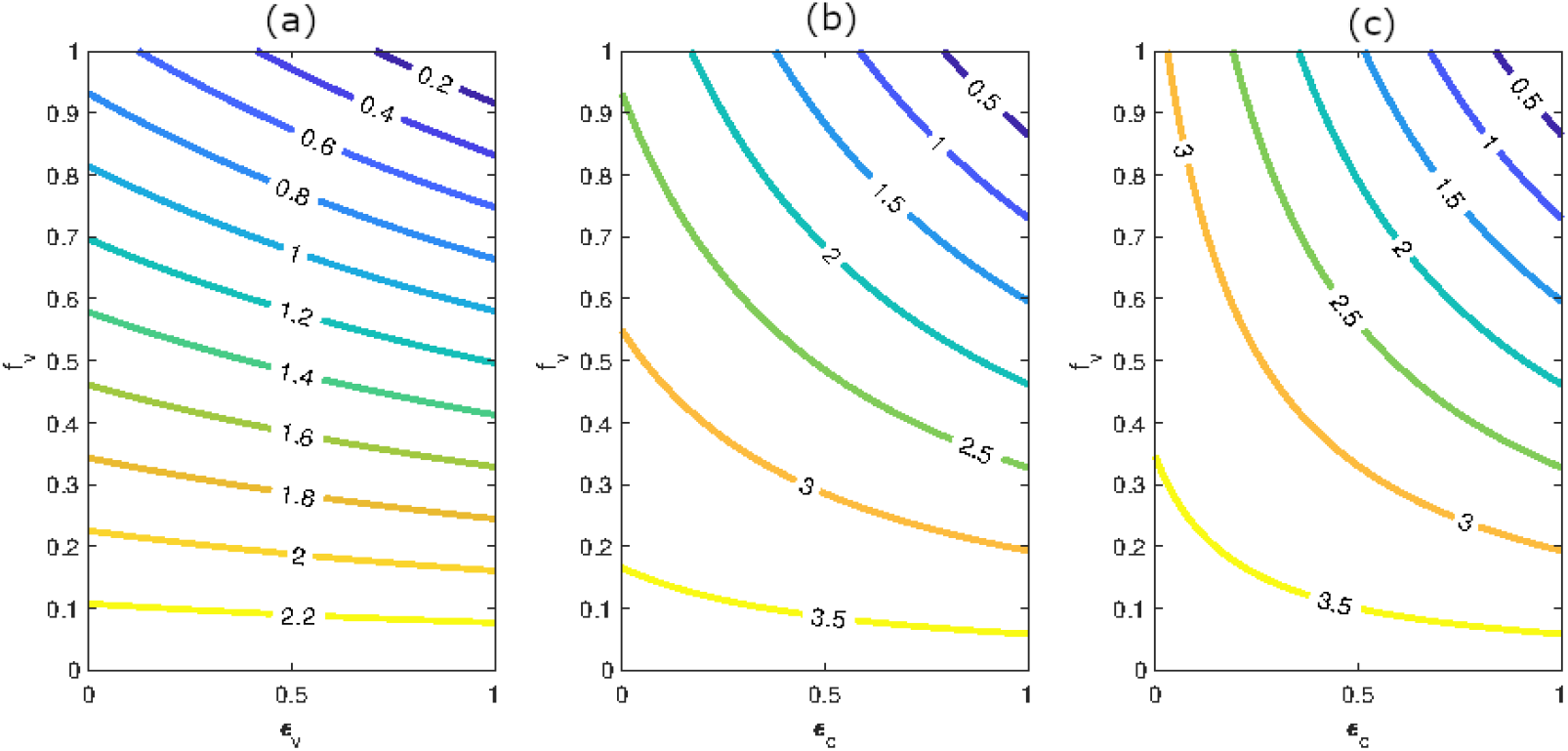
Contour plots of the vaccination reproduction numbers (*ℛ*_*vac*_) for the model (A-8), as a function of vaccine coverage at steady-state (*f*_*v*_) and vaccine efficacy against the wild-type strain (*ε*_*v*_) or the cross-protective efficacy offered by the vaccine against the variant strain *ε*_*c*_) for the US. The vaccination reproduction number for the (a) wild-type strain (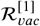, strain 1) with *β*_1_ = 0.24, (b) a variant strain that is 56% more transmissible than the wild-type strain (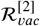, strain 2), and (c) a variant strain that is 100% more infectious than the wild-type strain (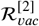, strain 2). Other parameter values used in this simulation are as given in Table A.9.

### 3.3. Parameter Sensitivity Analysis

The model (A-8) contains numerous parameters, and uncertainties arise in the estimated values of some of the parameters (e.g., those related to vaccination and the dynamics of the variant strains) used in the numerical simulations. In this section, global sensitivity analysis is carried out, using Latin Hypercube Sampling (LHS) and Partial Rank Correlation Coefficients (PRCC) (55; 11; 56), to determine the parameters that have the highest influence on the value of the chosen response function. Sensitivity analysis measures the extent that a response function changes with respect to variations in the input variables. For this study, the *vaccination reproduction number* for the variant strain 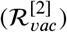 was used as the response function. One thousand bootstrap sample iterations were used for the LHS, and PRCC determined the correlation each individual parameter has on the response function 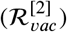. The baseline value of the contact rate for the variant strain, *β*_2_, was chosen as 0.2495. The baseline values for the additional fixed parameters are as given in Table A.9. Each parameter was assumed to be uniformly distributed and range from 50% to 150% of the baseline value. The PRCC values generated for the parameters in the expression of the response function, 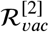, are tabulated in Table 2. It follows from this table that our studies identifies the following four parameters that play a more dominant role in the size of the vaccination reproduction number for the variant strain 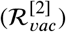:

**Table 2.** PRCC values for the parameters of the model (A-8), using the *vaccination reproduction number* of the variant strain 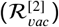 as the response function. Parameter values that most affect the dynamics of the model are highlighted in bold font.

| Parameter | Baseline | Range | PRCC | Parameter | Baseline | Range | PRCC |
| --- | --- | --- | --- | --- | --- | --- | --- |
| $\beta_2$ | 0.2495 | 0.1248-0.3744 | **0.8994** | $\phi_2$ | 0.02 | 0.010-0.030 | -0.0680 |
| $\epsilon_c$ | 0.50 | 0.25-0.75 | **-0.6215** | $\phi_{V2}$ | 0.01 | 0.005-0.015 | -0.0012 |
| $\eta_{P2}$ | 1.25 | 0.625-1.875 | 0.2942 | $f_v$ | 0.66 | 0.33-0.99 | **-0.8389** |
| $\eta_{PV2}$ | 1.25 | 0.625-1.875 | 0.2488 | $\mu$ | $3.5 \times 10^{-5}$ | $1.7 \times 10^{-5}$-<br>$5.20 \times 10^{-5}$ | -0.0255 |
| $\eta_{A2}$ | 0.75 | 0.375-1.125 | 0.3163 | $\gamma_{I2}$ | 0.10 | 0.05-0.15 | -0.4534 |
| $\eta_{AV2}$ | 0.75 | 0.375-1.125 | -0.0249 | $\gamma_{IV2}$ | 0.135 | 0.0675-0.2025 | -0.3822 |
| $\eta_{H2}$ | 0.25 | 0.125-0.375 | -0.0264 | $\gamma_{A2}$ | 0.20 | 0.10-0.30 | -0.1627 |
| $\eta_{HV2}$ | 0.25 | 0.125-0.375 | 0.0162 | $\gamma_{AV2}$ | 0.33 | 0.1650-0.4905 | -0.1105 |
| $\sigma_2$ | 0.40 | 0.20-0.60 | -0.0052 | $\gamma_{H2}$ | 0.0714 | 0.0357-0.1071 | -0.0821 |
| $\sigma_{V2}$ | 0.40 | 0.20-0.60 | 0.0333 | $\gamma_{HV2}$ | 0.083 | 0.0415-0.1245 | -0.0505 |
| $q$ | 0.60 | 0.30-0.90 | 0.2913 | $\delta_{I2}$ | 0.0015 | $7.5 \times 10^{-4}$-<br>0.0023 | 0.0091 |
| $q_v$ | 0.60 | 0.30-0.90 | 0.2272 | $\delta_{IV2}$ | 0.0001 | $5.0 \times 10^{-5}$-<br>$1.5 \times 10^{-4}$ | 0.0312 |
| $\xi_2$ | 0.40 | 0.20-0.60 | -0.3617 | $\delta_{H2}$ | 0.005 | 0.0025-0.0075 | -0.0232 |
| $\xi_{V2}$ | 0.40 | 0.20-0.60 | -0.2608 | $\delta_{HV2}$ | 0.0025 | 0.0013-0.0037 | 0.0427 |
| $\theta_c$ | 0.8 | 0.4-1.0 | **0.5699** | | | | |

i. the contact rate parameter for the transmission of the variant strain (*β*_2_);
ii. the cross protective efficacy of the variant strain (*ε*_*c*_);
iii. the steady-state proportion of fully-vaccinated individuals (*f*_*v*_); and
iv. the modification parameter accounting for the reduced infectiousness of fully-vaccinated individuals experiencing breakthrough infection (*θ*_*c*_).

In other words, the parameters *β*_2_, *ε*_*c*_, *f*_*v*_, and *θ*_*c*_ have the greatest influence on the *vaccination reproduction number* of the variant strain.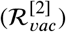. Hence, control measures against the variant strain (and, consequently, the disease in general) should be focused on these four parameters.

### 3.4. Numerical Simulations: Assessment of Vaccination Program

The model (A-8) is now simulated using the fixed parameters in Table A.9 to assess the population-level impact of the dominating emerging variant on the dynamics of COVID-19 in the US. The simulations are carried out for the period March 6, 2021 to March 6, 2024 for two levels of variant strain transmissibility (i.e., infectiousness) in relation to the wild-type strain: Scenario 1 assumes the variant strain is the B.1.1.7 (Alpha) variant (which is 56% more transmissible than the wild-type strain) (25)) and Scenario 2 assumes the variant strain is the Delta variant (which is 100% more transmissible than the wild-type strain) (19). The values of each model compartment on the last point of the fitting period on March 6, 2021 (see Section 2.2) were used as initial conditions to simulate projections of future COVID-19 dynamics in the US. The contact rate for the wild-type strain was chosen to be *β*_1_ =0.0805 per day from March 6, 2021 to March 30, 2021 (i.e., the result of fitting the post-third wave period, see Table A.9) and increased to *β*_1_ = 0.161 per day on April 1, 2021 to account for the relaxation of mask mandates and social distancing policies in various states (27; 3). Each Scenario is assessed for three levels of vaccination coverage: 50%, 66%, and 75% (i.e., *f*_*v*_ = 0.5, 0.66, and 0.75) and four levels of vaccination cross-protective efficacy against the variant strain (i.e., *ε*_*c*_ = 0.33, 0.5, 0.67, and 0.75). We assume, in these simulations, that vaccinated individuals received both doses of the Pfizer/Moderna vaccine (these vaccines have protective efficacy *ε*_*v*_= 0.94).

For mathematical tractability, it is assumed that recovered individuals do not acquire further SARS-CoV-2 infection. This assumption is consistent with other COVID-19 modeling studies (38; 40; 59; 58; 46; 29). Furthermore, since less than 8% of fully-vaccinated individuals in the US have received the Johnson & Johnson vaccine (13), simulations under Pfizer/Moderna vaccine administration can better estimate the future course of the pandemic.

#### 3.4.1. Scenario 1: Variant is 56% more transmissible than the wild-type strain

Data from Korber et al. (50) suggests that the B.1.1.7. (Alpha) variant is 56% more transmissible than the wild-type strain in the US. Here, we simulate the dynamics of the two strains (wild-type strain and the Alpha variant) for various levels of the coverage level of fully-vaccinated individuals (namely 50%, 66%, 75%) and cross-protective vaccination efficacy (*ε*_*c*_ = 0.33, 0.5, 0.67, 0.75) offered by the Pfizer/Moderna vaccines. The cumulative COVID-19 mortality and cases, generated using the model for Scenario 1, are tabulated in Table 3. This table shows that if 66% of the US population becomes fully-vaccinated with the Pfizer/Moderna vaccines, then under vaccination cross efficacy levels between 33% and 75% (i.e., *ε*_*c*_ ranges from 0.33 to 0.75), COVID-19 related deaths can be reduced between 27% and 44%, and cumulative cases can be reduced between 22% and 46%, when compared to the case where only 50% of the US population is fully-vaccinated. Furthermore, if as high as 75% of the US population becomes fully-vaccinated, then the reduction of COVID-19 related deaths and cases can increase to as high as 51% and 55%, respectively (when compared to the case when only 50% of the US population is fully-vaccinated).

**Table 3.** Simulations of the model (A-8), showing the cumulative COVID-19 mortality and infections under Scenario 1, for varying levels of cross-protective efficacy of the vaccine against the variant strain (*ε*_*c*_) and vaccination coverage (*f*_*v*_), as of March 6, 2024 in the US. Scenario 1 assumes vaccinated individuals receive two doses of the Pfizer or Moderna vaccine (*ε*_*v*_ = 0.94) and that the variant strain is 56% more transmissible than the wild-type strain.

| $\epsilon_c$ | $f_v$ | Cumulative Deaths ($\times 10^5$) | | Cumulative New Infections ($\times 10^6$) | |
| --- | --- | --- | --- | --- | --- |
|  |  | Both Strains | Variant | Both Strains | Variant |
| 0.33 | 0.5 | 13.17 | 7.60 | 119.33 | 73.79 |
|  | 0.66 | 10.00 | 4.44 | 91.72 | 46.28 |
|  | 0.75 | 8.06 | 2.50 | 72.80 | 27.3 |
| 0.5 | 0.5 | 11.42 | 5.86 | 100.84 | 55.29 |
|  | 0.66 | 6.46 | 0.90 | 54.85 | 9.34 |
|  | 0.75 | 5.57 | 0.12 | 45.64 | 0.124 |
| 0.67 | 0.5 | 9.33 | 3.77 | 79.8 | 34.25 |
|  | 0.66 | 5.56 | 0.0047 | 45.55 | 0.043 |
|  | 0.75 | 5.56 | 0.0030 | 45.54 | 0.030 |
| 0.75 | 0.5 | 8.25 | 2.69 | 69.48 | 23.94 |
|  | 0.66 | 5.56 | 0.0026 | 45.52 | 0.023 |
|  | 0.75 | 5.56 | 0.0020 | 45.52 | 0.020 |

Table 4 tabulates the values and occurrence of the peak daily COVID-19 related deaths and new infections generated using the model. It can be seen from Table 4 that a future COVID-19 wave or surge can be averted if at least 66% of the US population is fully-vaccinated with either the Pfizer or Moderna vaccine, and the cross-protective efficacy the vaccine offers against the Alpha variant is at least 67%. Even if the cross-protective efficacy level is as low as 50%, a future pandemic wave can be avoided if at least 75% of the US populace is fully-vaccinated with either of the aforementioned vaccines.

**Table 4.** Simulations of the model (A-8), showing the peak of daily COVID-19 related deaths and infections from March 6, 2021 until March 6, 2024, under Scenario 1, for various levels of cross-protective vaccination efficacy against the variant strain (*ε*_*c*_) and vaccination coverage (*f*_*v*_). Scenario 1 assumes vaccinated individuals receive two doses of the Pfizer or Moderna vaccines (*ε*_*v*_ = 0.94) and that the variant strain is 56% more as infectious than the wild-type strain. A dashed line “-” indicates that the simulation did not produce a new peak of infections or deaths during the course of simulation.

| $\epsilon_c$ | $f_v$ | Peak of Daily Deaths ($\times 10^3$) | | Peak of Daily New Infections ($\times 10^5$) | |
| --- | --- | --- | --- | --- | --- |
|  |  | Date | Amount | Date | Amount |
| 0.33 | 0.5 | Feb 10, 2022 | 6.17 | Jan 30, 2022 | 6.20 |
|  | 0.66 | June 22, 2022 | 2.03 | June 12, 2022 | 4.44 |
|  | 0.75 | Jan 10, 2023 | 0.63 | Dec 20, 2022 | 0.69 |
| 0.5 | 0.5 | May 7, 2022 | 3.51 | April 25, 2022 | 3.38 |
|  | 0.66 | Jan 18, 2024 | 0.24 | Jan 4, 2024 | 0.24 |
|  | 0.75 | — | — | — | — |
| 0.67 | 0.5 | Oct 30, 2022 | 1.43 | Oct 19, 2022 | 1.31 |
|  | 0.66 | — | — | — | — |
|  | 0.75 | — | — | — | — |
| 0.75 | 0.5 | April 12, 2023 | 0.77 | March 27, 2023 | 0.69 |
|  | 0.66 | — | — | — | — |
|  | 0.75 | — | — | — | — |

#### 3.4.2. Scenario 2: Variant is 100% more transmissible than the wild-type strain

Here, we simulated the case where the wild-type strain is co-circulating with the Delta variant (which is 100% more transmissible than the wild-type strain). For these simulations, the Pfizer and Moderna vaccines are used. Cumulative COVID-19 related deaths and cases for Scenario 2 are tabulated in Table 5. This table shows that under 50% full-vaccination coverage and the same levels of cross-efficacy protection against the variant strain (i.e., *ε*_*c*_ ranging from 0.33 to 0.75), a 100% more infectious variant may result in 21% to 50% more COVID-19 related deaths and between 31% to 54% more cases, when compared to the 50% vaccination coverage level with the Alpha variant (i.e., a variant that is only 56% more infectious than the wild-type strain). Thus, the results in this table show a higher burden of the COVID-19 will be recorded in the US if the wild-type strain is co-circulating with the Delta strain, as against co-circulation with the Alpha variant (as expected). This table further shows that if the vaccination coverage is increased to 75%, then the COVID-19 related deaths and cases can be reduced by 24-56% and 20-58%, respectively, in comparison to only having 50% full-vaccination coverage, under this more infectious variant.

**Table 5.** Cumulative COVID-19 related deaths and infections simulated by the model (A-8) under Scenario 2 for varying levels of cross-protective vaccination efficacy against the variant strain (*ε*_*c*_) and vaccination coverage (*f*_*v*_) as of March 6, 2024 in the US. Scenario 2 assumes that vaccinated individuals receive two doses of the Pfizer or Moderna (*ε* _*v*_= 0.94) and that the variant strain is 100% more infectious than the wild-type strain.

| $\epsilon_c$ | $f_v$ | Cumulative Deaths ($\times 10^5$) | | Cumulative New Infections ($\times 10^6$) | |
| --- | --- | --- | --- | --- | --- |
|  |  | Both Strains | Variant | Both Strains | Variant |
| 0.33 | 0.5 | 16.63 | 11.06 | 153.78 | 108.23 |
|  | 0.66 | 14.05 | 8.49 | 135.06 | 89.56 |
|  | 0.75 | 12.61 | 7.05 | 123.00 | 77.50 |
| 0.5 | 0.5 | 15.23 | 9.67 | 138.00 | 92.47 |
|  | 0.66 | 11.77 | 6.21 | 109.06 | 63.56 |
|  | 0.75 | 9.64 | 4.09 | 89.00 | 43.51 |
| 0.67 | 0.5 | 13.41 | 7.84 | 117.9 | 72.36 |
|  | 0.66 | 8.82 | 3.26 | 77.18 | 31.67 |
|  | 0.75 | 5.95 | 0.39 | 49.46 | 3.95 |
| 0.75 | 0.5 | 12.39 | 6.83 | 107.12 | 61.57 |
|  | 0.66 | 7.26 | 1.70 | 61.54 | 16.01 |
|  | 0.75 | 5.62 | 0.07 | 46.18 | 0.66 |

Figure 5 shows the simulations of Scenario 2 when 75% of the US population is fully vaccinated with the Pfizer or Moderna vaccines. The values and occurrence of the peak daily COVID-19 related deaths and new infections are included in Table 6. Table 6 shows that under this more infectious variant, a future pandemic wave can only be avoided if the US attains a full-vaccination coverage of 75% under the Pfizer and Moderna vaccines, and the cross-efficacy provided by the aforementioned vaccines against the Delta variant is at least 67%.

**Table 6.** The peak daily COVID-19 related deaths and infections from March 6, 2021 to March 6, 2024 simulated by the model (A-8) under Scenario 2 varying levels of cross-protective vaccination efficacy against the variant strain (*ε*_*c*_) and vaccination coverage (*f*_*v*_). Scenario 2 assumes vaccination individuals receive two doses of the Pfizer or Moderna vaccines (*ε*_*v*_ = 0.94) and that the variant strain is 100% more infectious than the wild-type strain. A dashed line “-” indicates that the simulation did not produce a new peak of infections or deaths during the course of simulation.

| $\epsilon_c$ | $f_v$ | Peak Daily Deaths ($\times 10^3$) | | Peak Daily New Infections ($\times 10^5$) | |
| --- | --- | --- | --- | --- | --- |
|  |  | Date | Amount | Date | Amount |
| 0.33 | 0.5 | Sept 20, 2021 | 15.86 | Sept 10, 2021 | 17.16 |
|  | 0.66 | Oct 10, 2021 | 9.30 | Sept 30, 2021 | 10.36 |
|  | 0.75 | Oct 20, 2021 | 6.37 | Oct 11, 2021 | 7.26 |
| 0.5 | 0.5 | Oct 13, 2021 | 11.78 | Oct 3, 2021 | 12.15 |
|  | 0.66 | Nov 30, 2021 | 4.72 | Nov 19, 2021 | 4.96 |
|  | 0.75 | Jan 11, 2022 | 1.98 | Jan 1, 2022 | 2.13 |
| 0.67 | 0.5 | Nov 17, 2021 | 7.70 | Nov 6, 2021 | 7.48 |
|  | 0.66 | Apr 3, 2022 | 1.26 | Mar 24, 2022 | 1.23 |
|  | 0.75 | — | — | — | — |
| 0.75 | 0.5 | Dec 9, 2021 | 5.95 | Nov 27, 2021 | 5.60 |
|  | 0.66 | Sept 20, 2022 | 0.35 | Aug 20, 2022 | 0.33 |
|  | 0.75 | — | — | — | — |

**Figure 4:**
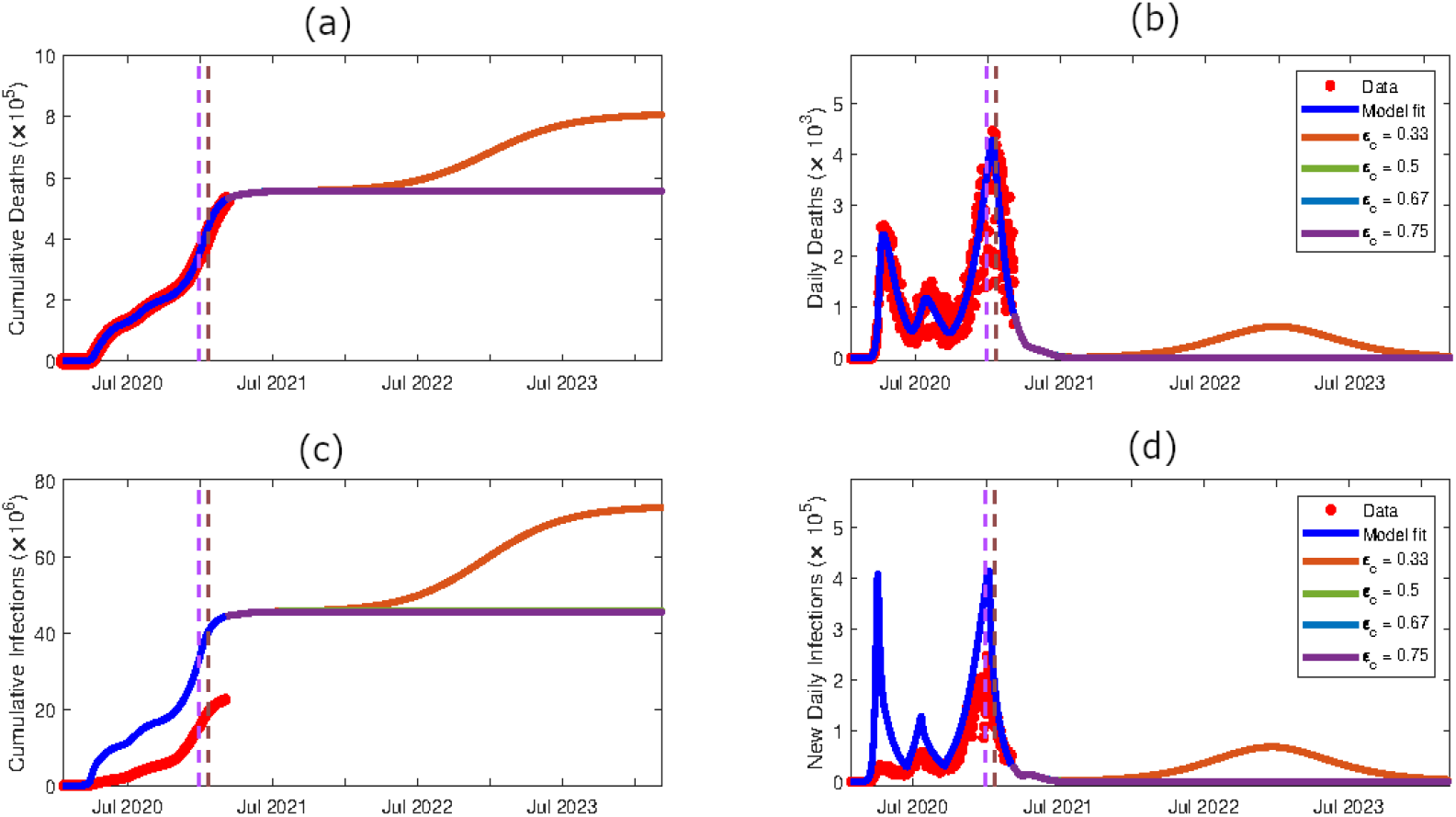
Simulations of the model (A-8), showing COVID-19 projection for the period from January 22, 2020 to March 6, 2024, for Scenario 1 in the US. In these simulations, the Pfizer/Moderna vaccine (with *ε*_*v*_ = 0.94) is used, together with 75% vaccination coverage and the variant strain is 56% more transmissible than the wild-type strain (i.e., we consider the Alpha variant in these simulations). The values of the remaining parameters of the model are as given in Table A.9. (a) cumulative COVID-19 mortality, (b) daily COVID-19 mortality, (c) cumulative COVID-19 infections, (d) new daily COVID-19 infections. The red dots represent the reported data points from (21), the dark blue curve indicates the model fit from Section 2.2, the red curve shows the projections assuming low cross-efficacy of the variant strain (strain 2) from the vaccine (*ε*_*c*_ = 0.33), the green curve shows projections assuming a moderate amount of cross-protective efficacy (*ε*_*c*_ = 0.5), the light blue curve shows projections assuming a moderately-high amount of cross-protective efficacy (*ε*_*c*_ = 0.67) and the purple curve shows the projections assuming a high amount of cross-protective efficacy (*ε*_*c*_ = 0.75). The purple dashed vertical line for each panel indicates the time the variant strain was introduced into the population (December 29, 2020), and the brown dashed vertical line represents the time vaccination was introduced into the model (January 22, 2021).

**Figure 5:**
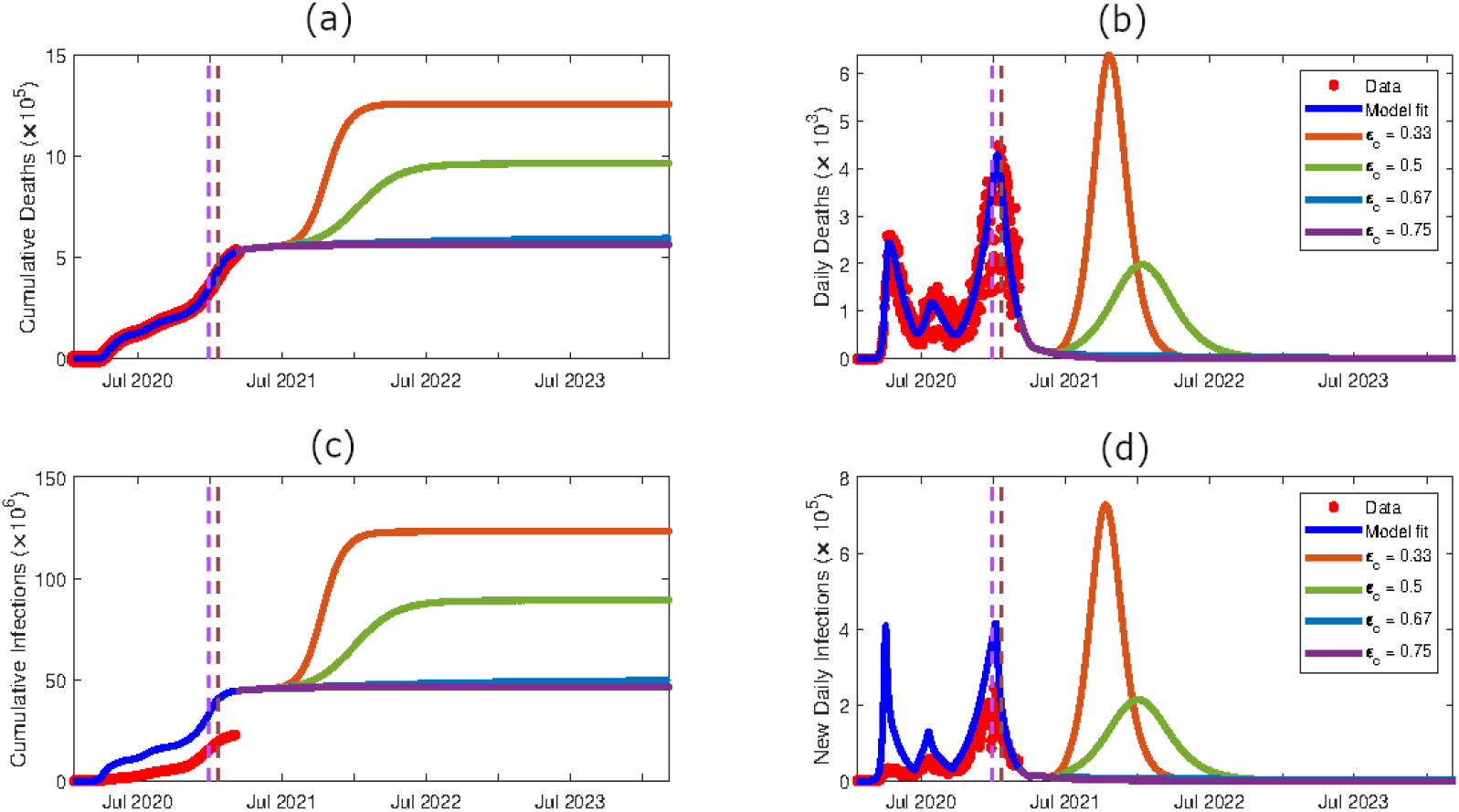
Simulation projections of model (A-8) from January 22, 2020 to March 6, 2024 for Scenario 2 in the U.S: Pfizer or Moderna vaccine (*ε*_*v*_ = 0.94) and 75% vaccination coverage with variant strain (strain 2) 100% more infectious than the wild-type strain (strain 1). Additional parameter values are included in Table A.9. (a) cumulative COVID-19 mortality, (b) daily COVID-19 mortality, (c) cumulative COVID-19 infections, (d) new daily COVID-19 infections. The red dots represent the reported data points, the dark blue curve indicates the model fit from Section 2.2, the red curve shows the projections assuming the vaccines produce a low level of cross-efficacy of the variant strain (strain 2) from the vaccine (*ε*_*c*_ = 0.33), the green curve shows projections assuming a moderate amount of cross-protective efficacy (*ε*_*c*_ = 0.5), the light blue curve shows projections assuming a moderately-high amount of cross-protective efficacy (*ε*_*c*_ = 0.67) and the purple curve shows the projections assuming a moderately-high amount of cross-protective efficacy (*ε*_*c*_ = 0.75). The purple dashed vertical line for each panel indicates the time when variant cases were introduced into the US population (December 29, 2020), and the brown dashed vertical line represents the time vaccination was introduced into the model (January 22, 2021).

## 4. Discussion & Conclusions

Since the emergence of COVID-19 in Wuhan City, China on December 31, 2019 (82; 86), the pandemic has induced severe public health and socio-economic burden on an unprecedented scale. Fortunately, several safe and effective vaccines were developed in record time to combat the virus. Specifically, two mRNA-based (Pfizer and Moderna) and an adenovirus-based (Johnson & Johnson) vaccines were given emergency use authorization (EUA) in the US from as early as December 2020 (the Johnson & Johnson vaccine received EUA in March 2021). Nearly 50% of the US population has attained full-vaccination status as of August 3, 2021 (13). However, the recent emergence of SARS-CoV-2 variants have posed major public health concerns in the US and globally. In particular, the B.1.1.7 (Alpha) and the B.1.617.2 (Delta) variants, that first emerged in the UK and India, respectively, have been circulating in many parts of the world, including the US (19). Current data and studies show strong evidence that these variants are more easily transmissible, have higher rates of hospitalization and increased mortality rates, in comparison to the wild-type strain (45). One of the greatest challenges associated with COVID-19 vaccination programs is the uncertainty of the level of cross-protection from currently-available COVID-19 vaccines offer against the variant strains.

Numerous mathematical modeling studies, using varying modeling types and paradigms, have been conducted to provide insight into the transmission dynamics and control of COVID-19 in different populations. The modeling types used include statistical (48; 66; 64), mechanistic/deterministic (58; 38; 40), stochastic (42; 44; 51), network (32; 68; 85), and agent-based (23; 31). The current study uses a deterministic system of 26 nonlinear differential equations to assess the population-level impact of the cross-protective efficacy of vaccination on SARS-CoV-2 variant strains. We used known parameter values for the wild-type strain and open-access COVID-19 death data to estimate the contact rate for infection transmission throughout the course of the pandemic in the United States. The disease-free equilibrium of the autonomous version (where the contact rate is assumed to be constant) of the resulting two-strain model (A-8), which incorporates the vaccination of susceptible and naturally recovered individuals, is shown to be asymptotically-stable whenever the associated *vaccination reproduction number* of the model (denoted by *ℛ*_*vac*_) is less than unity. The threshold quantity *ℛ*_*vac*_ measures the average number of new SARS-CoV-2 cases generated by a typical infected individual introduced into a population that is partially-protected (*via* vaccination and natural recovery from infection). An explicit expression for the vaccine-derived herd immunity threshold (denoted by 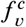) is derived. The herd immunity threshold is a measure of the minimum fraction of susceptible individuals in the population that need to be immunized to ensure the protection of the individuals that cannot be vaccinated, and, consequently, leading to the suppression or halting of the COVID-19 pandemic.

Our study shows that if only the wild-type SARS-Cov-2 strain is circulating in the US, then at least 59% of the populace needs to be fully-vaccinated (with either the Pfizer or Moderna vaccine, each with estimated protective efficacy of at least 94%) to achieve the vaccine-derived herd immunity. In other words, if only the original wild-type SARS-CoV-2 strain is circulating in the US (which was the case prior to the emergence of the SARS-CoV-2 variants in the US around December 2020 (19)), then at least 6 in every 10 people living in the US would need to be fully-vaccinated with the Pfizer/Moderna vaccines to achieve the vaccine-derived herd immunity. This herd value of vaccine-derived herd immunity, which is reasonably attainable in the US (13), is consistent with the herd immunity threshold derived in previous studies, such as those in (40; 38).

However, if the wild-type strain is co-circulating with the Alpha variant (which is 56% more transmissible than the wild-type strain), our study shows that the minimum threshold for achieving the vaccine-derived herd immunity (using either the Pfizer or Moderna vaccine) increases to 76%. The value increases to 82% if the wild-type is co-circulating with the Delta variant (which is 100% more transmissible than the wild-type strain). In other words, the emergence and circulation of the highly-transmissible Alpha and Delta variants contribute in raising the minimum value of the vaccine-derived herd immunity threshold needed to effectively control, and mitigate the burden of, COVID-19 pandemic in the US. While achieving 60% vaccination coverage against the COVID-19 pandemic if only the wild-strain is circulating is certainly realistically-attainable in the US, achieving 76% or 82% coverage, if any of the two variants are circulating, is undoubtedly a tall order, for the numerous factors that contributed to the concerning level of vaccine hesitancy in the US (13).

By implementing a global sensitivity analysis of the model, using the vaccination reproduction for the variant strain as the response function, this study identifies four parameters that have the highest influence on the SARS-CoV-2 variants (hence, of the COVID-19 pandemic) in the US. The identified parameters are the effective contact rate for the transmission of the SARS-CoV-2 variant strain contact rate (*β*_2_), the cross-protective efficacy offered by the Pfizer/Moderna vaccine against the variant strain (*ε*_*c*_), the coverage of fully-vaccinated individuals in the US (*f*_*v*_), and the relative infectiousness of fully-vaccinated infectious individuals, in comparison to the infectiousness of unvaccinated infectious individuals (*θ*_*c*_).

We carried out extensive numerical simulations of the two-strain model to assess the population-level impact of various SARS-CoV-2 dynamic scenarios, based on varying the vaccination efficacy against the wild-type strain (*ε*_*v*_), the vaccination cross-protective efficacy (*ε*_*c*_) and the coverage of the fully-vaccinated population (*f*_*v*_) for the case where the wild-type strain is co-circulating with the Alpha or the Delta variant. These simulations show, as expected, that wide-scale vaccination is crucial to mitigating the burden and/or eliminating the COVID-19 pandemic in the US. For instance, for the Alpha variant, which was shown to be 56% more infectious than the wild-type strain (25), we showed that increasing the coverage of fully-vaccinated individuals from the current 50% for the US to 75% can reduce anywhere from 33-51% of the cumulative mortality and 35-55% of cumulative cases (in comparison to when the vaccination coverage is 50%). For a SARS-CoV-2 variant that is more infectious than the Alpha variant, such as the Delta variant (which is 100% more transmissible than the wild-type strain), our simulations show that increasing the coverage of fully-vaccinated individuals from 50% to 75% could reduce anywhere from 24-56% of COVID-19 related deaths and 20-57% of cases, compared to the case with only 50% vaccination coverage.

Our study clearly shows that the vaccination coverage, cross-protective vaccination efficacy against variant strains, and the relative infectiousness of the variant strain, in comparison to the wild-type strain all greatly influence the dynamics of the COVID-19 pandemic in the US. Specifically, both the cross-protective vaccination efficacy against the variant and the vaccination coverage shift the peak of daily cases and deaths forward in time. This shifting increases with increasing values of the cross-protective efficacy and vaccination coverage. Our simulations show that, in general, the peak of daily deaths is expected to occur approximately 10 to 16 days after the peak of daily infections occurs.

One limitation of our study is the fact that our numerical simulations are based on the assumption that only the Pfizer or the Moderna vaccines are used in the population. In reality, three vaccines are being used in the US (the aforementioned two and the Johnson & Johnson vaccine), each with a presumably different level of cross-protective efficacy against the variant strains circulating in the population. Further, each of the three vaccines may differ in terms of their therapeutic properties (such as accelerating recovery, reducing risk of severe disease, hospitalization and deaths in breakthrough infections). Another limitation of our study is that the model is formulated to account for a single variant (i.e., we lumped all variants into a single dominant one). Although this assumption was necessarily made to enable mathematical tractability, extending the modeling framework to allow for the assessment of the dynamics of multiple SARS-CoV-2 variant strains (as is the case at the current time in the US, where both Alpha and Delta variants are co-circulating, but with the Delta clearly the prohibitive dominant one) would be quite useful. Since recent studies have shown that naturally-acquired immunity is less likely to produce an immune response to the existing variants than vaccination (62; 36), another future extension of the modeling framework we developed in this study is to include the possibility that individuals who recovered from infection from the wild-type strain (i.e., those in the *R*_1_ class in our model) could acquire infection with the variant strain without losing their natural immunity against the wild-type strain.

The Alpha (B.1.1.7) variant was the most dominant SARS-CoV-2 variant circulating in the US throughout April and May 2021 (15; 20), and was estimated to be 56% more transmissible than the wild-type strain (25). The numerical simulations we conducted in this study showed that if a moderate level of vaccination coverage (≥ 66% of the US population) and moderately-high level of cross-protective vaccination efficacy is provided by the Pfizer and Moderna vaccines (*ε*_*c*_ ≥ 0.67), then a surge of variant-induced COVID-19 infections and mortality will not occur (under the assumption that both the vaccine-derived and naturally-acquired immunity lasts at least two years). A recent study showed that cross-protective efficacy is as high as 93% against the Alpha variant when two doses of the Pfizer vaccine are administered, but drops to 33% when only one dose is received (37). Thus, strictly following the two-dose regimen for the Pfizer and Moderna vaccines is essential to significantly reduce community transmission of COVID-19. The insights generated from this study align with the findings from other recent modeling studies for the dynamics of wild-type and variant strains of SARS-Cov-2 (also using SEIR-type modeling paradigm) (8; 34).

Many US states relaxed control and mitigation measures against COVID-19, such as face mask mandates, once COVID-19 vaccination had become more widely available (27; 3). Unfortunately, vaccination rates have since slowed down, since April 2021 (13), and vaccinated individuals can still experience breakthrough cases that are infectious to others (14). Since June 2021, the Delta variant has become the dominant SARS-Cov-2 variant in the US (20), and is known to be more infectious than the Alpha variant (5). For the Delta variant (which is 100% more transmissible than the wild-type strain), our simulations show that the COVID-19 pandemic can be effectively controlled in the US if 75% of the US population is fully-vaccinated (with either the Pfizer or Moderna vaccine) and the vaccine offers at least 67% cross-protective efficacy against the Delta variant. Thankfully, it was shown that the full recommended dosage of the Pfizer vaccine is 88% effective against the Delta variant (37). However, if the vaccination coverage is lower, and a highly-transmissible variant (such as Delta) is co-circulating, the US still risks experiencing a severe COVID-19 wave in the future. As of August 2021, the US has experienced increased COVID-19 related cases and deaths (53), with 97% and 99% of the COVID-19 related hospitalizations and deaths exclusively occurring in unvaccinated infected individuals, respectively (65). Some US cities have since responded by resuming NPI (nonpharmaceutical intervention) mandates, such as mask wearing and social-distancing, in line with the new CDC recommendations (47). Thus, it is critical to continue the practice of following NPIs, and encourage all eligible individuals to receive the full two-dose regimen of the Pfizer or Moderna vaccines. In summary, our study demonstrated that the prospect for the effective control and/or elimination of the SARS-CoV-2 variants (hence, the COVID-19 pandemic) in the US is promising provided the coverage of the fully-vaccinated individuals, and the level of cross-protection the vaccine offers against the SARS-CoV-2 variant strains, are high enough.

## Data Availability

Data used for simulations are publicly available at Johns Hopkins University COVID-19 repository

## Acknowledgments

One of the authors (ABG) acknowledge the support, in part, of the Simons Foundation (Award #585022) and the National Science Foundation (grant number: DMS-2052363). The authors are grateful to the anonymous reviewers for their constructive comments.

## Appendix A

### Equations of the Mathematical Model

The two-strain, two-group vaccination model for the COVID-19 transmission dynamics and control in the US is given by the following deterministic system of nonlinear differential equations:

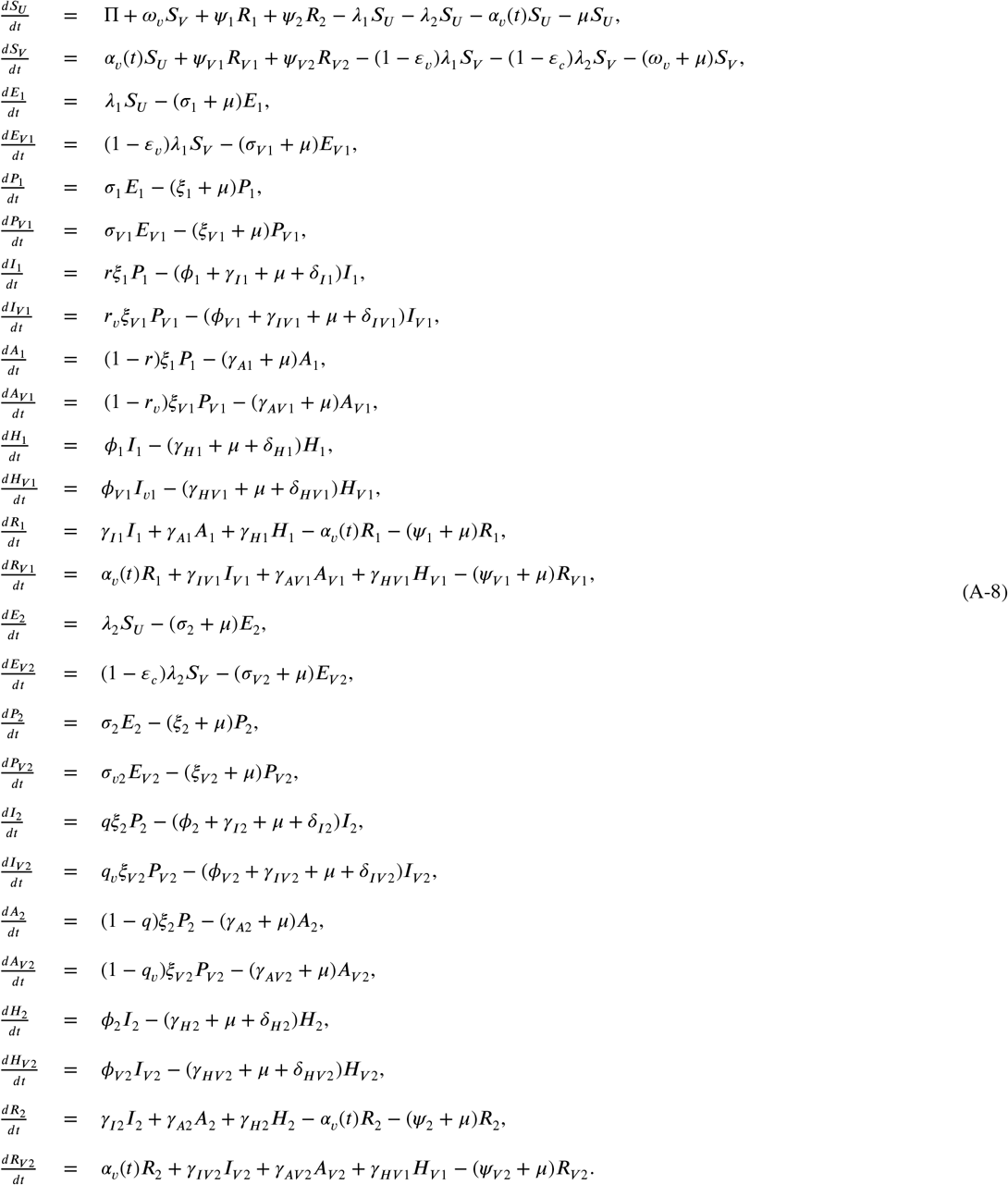

### Description of Parameters of the Model

The parameter II is the recruitment rate (via birth or immigration) of individuals into the population. Individuals are vaccinated at a time-dependent rate *α*_*v*_(*t*). The vaccine is assumed to produce protective efficacy 0 < *ε*_*v*_ < 1 against strain 1, potential cross-protective efficacy 0 ≤ *ε*_*c*_ < 1 against strain 2, and wane at rate *ω*_*v*_. Natural death occurs in all epidemiological classes at rate *µ*. Susceptible individuals acquire infection with strain 1 at a rate *λ*_1_ or with strain 2 at a rate *λ*_2_, where

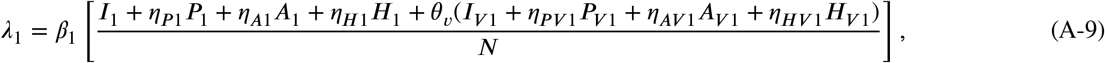

and,

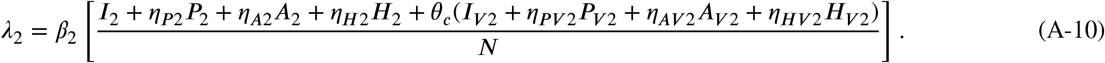

**Table A.7.** Description of variables of model (A-8) where (*i* = 1, 2) indicates the strain of infection.

| Variable ($i = 1, 2$) | Interpretation |
| --- | --- |
| $S_U (S_V)$ | Population of unvaccinated (vaccinated) susceptible individuals |
| $E_i (E_{Vi})$ | Population of unvaccinated (vaccinated) individuals exposed to (latently infected with) strain $i$ |
| $P_i (P_{Vi})$ | Population of unvaccinated (vaccinated) pre-symptomatic infectious individuals infected with strain $i$ |
| $I_i (I_{Vi})$ | Population of unvaccinated (vaccinated) symptomatic infectious individuals infected with strain $i$ |
| $A_i (A_{Vi})$ | Population of unvaccinated (vaccinated) asymptomatic infectious individuals infected with strain $i$ |
| $H_i (H_{Vi})$ | Population of unvaccinated (vaccinated) hospitalized infectious individuals infected with strain $i$ |
| $R_i (R_{Vi})$ | Population of unvaccinated (vaccinated) individuals who naturally recovered from strain $i$ infection |

In (A-9) and (A-10), *β*_*i*_ (with = 1, 2) is the effective contact rate for strain, *i, η*_*Pi*_, (*η*_*Pvi*_), *η*_*Ai*_ (*η*_*Avi*_) and *η*_*Hi*_ (*η*_*Hvi*_) are, respectively, the modification parameters accounting for the assumed increase or reduction of contact rates of unvaccinated (vaccinated) pre-symptomatic, asymptomatic, and hospitalized individuals with strain, compared to symptomatic infectious individuals with strain. The modification parameter 0 ≤ *θ*_*v*_ ≤ 1 represents the assumed reduced infectiousness of individuals vaccinated against strain 1 (in comparison to unvaccinated infectious individuals with strain 1), while (0 ≤ *θ*_*c*_ ≤ 1) accounts for the assumed reduced infectiousness of strain 2 induced by the cross-protective of vaccination against strain 1.

Unvaccinated (vaccinated) individuals in the exposed classes (strain 1 or strain 2) progress to their respective strain’s pre-symptomatic class at rate *σ*_*i*_ (*σ*_*vi*_) where = {1, 2} indicates the strain of infection. Non-vaccinated (vaccinated) individuals transition out of the pre-symptomatic classes at rate *ξ*_*i*_ (*ξ*_*vi*_). For strain 1, fraction 0 < *r* (*r*_*v*_) < 1 of individuals develop symptoms while (1 − *r*) ((1 − *r*_*v*_)) individuals remain asymptomatic infectious. The fraction of unvaccinated (vaccinated) individuals infected with strain 2 who develop symptoms is 0 < *q* (*q*_*v*_) < 1. Non-vaccinated (vaccinated) symptomatic infectious individuals recover at rate *σ*_*Ii*_ (*σ*_*Ivi*_), become hospitalized at rate *ϕ*_*i*_ (*ϕ*_*vi*_), and die from diseased-induced death at rate *δ*_*Ii*_ (*δ*_*Ivi*_). Non-vaccinated (vaccinated) asymptomatic infectious individuals recover at rate *σ*_*Ai*_ (*σ*_*Avi*_). Non-vaccinated (vaccinated) Hospitalized individuals recover at rate *σ*_*Hi*_ (*σ*_*Hvi*_) and die from disease-induced death rate *δ*_*Hi*_ (*δ*_*Hvi*_). Non-vaccinated (vaccinated) individuals lose their naturally-acquired disease-induced immunity at rate *ψ*_*i*_ (*ψ*_*vi*_).

### Table of State Variables and Parameters of the Model

The state variables and parameters of the model are described in Tables A.7 and A.8, respectively.

### Values of Fixed Parameters

**Table A.8.** Description of parameters of model (A-8) where (*i* = {1, 2}) indicates the strain of infection.

| Parameter ($i = 1, 2$) | Interpretation |
| --- | --- |
| $\Pi$ | Recruitment rate into the population via birth or immigration |
| $\beta_i$ | Infectious contact rate of individuals infected with strain $i$ |
| $\eta_{Pi} (\eta_{Ai}, \eta_{Hi})$ | Modification parameter to account for relative infectiousness of unvaccinated pre-symptomatic (asymptomatic, hospitalized) individuals infected with strain $i$ |
| $\eta_{PVi} (\eta_{AVi}, \eta_{HVi})$ | Modification parameter to account for relative infectiousness of vaccinated pre-symptomatic (asymptomatic, hospitalized) individuals infected with strain $i$ |
| $0 \leq \theta_v (\theta_c) \leq 1$ | Modification parameter to account for reduced infectiousness of vaccinated individuals infected with strain 1 (strain 2) |
| $\alpha_v(t)$ | *Per capita* vaccination rate |
| $\omega_v$ | Waning rate of vaccine |
| $\psi_i (\psi_{Vi})$ | Rate of loss of disease-acquired immunity for unvaccinated (vaccinated) individuals infected with strain $i$ |
| $\mu$ | Natural death rate |
| $0 \leq \varepsilon_v \leq 1$ | Vaccination efficacy against the wild-type strain (strain 1) |
| $0 \leq \varepsilon_c \leq 1$ | Vaccination cross-protective efficacy against the variant strain (strain 2) |
| $\xi_i (\xi_{Vi})$ | Progression rate from unvaccinated (vaccinated) exposed (latently infected) individuals infected with strain $i$ to pre-symptomatic class |
| $0 < r (r_v) < 1$ | Fraction of unvaccinated (vaccinated) infectious individuals infected with strain 1 who develop clinical symptoms of COVID-19 |
| $0 < q (q_v) < 1$ | Fraction of unvaccinated (vaccinated) infectious individuals infected with strain 2 who develop symptoms |
| $\phi_i (\phi_{Vi})$ | Hospitalization rate of unvaccinated (vaccinated) symptomatic infectious individuals infected with strain $i$ |
| $\gamma_{Ii} (\gamma_{Ai}, \gamma_{Hi})$ | Recovery rate of unvaccinated symptomatic infectious (asymptomatic infectious, hospitalized) individuals infected with strain $i$ |
| $\gamma_{IVi} (\gamma_{AVi}, \gamma_{HVi})$ | Recovery rate of vaccinated symptomatic infectious (asymptomatic infectious, hospitalized) individuals infected with strain $i$ |
| $\delta_{Ii} (\delta_{IVi})$ | Disease-induced death rate of unvaccinated (vaccinated) symptomatic infectious individuals infected with strain $i$ |
| $\delta_{Hi} (\delta_{HVi})$ | Disease-induced death rate of unvaccinated (vaccinated) hospitalized individuals infected with strain $i$ |

## Appendix B

### Computation of *ℛ*_*vac*_

The associated next generation matrices of the model (A-8) are given by:

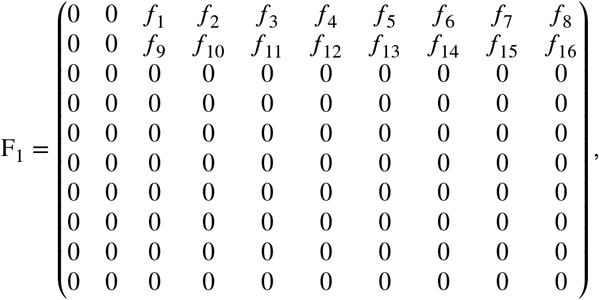

where (noting that 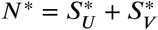, with 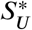 and 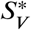 defined in Section 3):

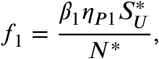

**Table A.9.** Fixed parameters of the model (A-8) for data fitting and simulations. The units of all rate parameters is *per* day

| Parameter | Value | Source | Parameter | Value | Source | Parameter | Value | Source |
| --- | --- | --- | --- | --- | --- | --- | --- | --- |
| $\Pi$ | 12,000 | (40) | $\mu$ | $3.5 \times 10^{-5}$ | (40) | $\gamma_{I2}$ | 0.1 | (59; 58)* |
| $\eta_{P1}$ | 1.25 | (40) | $\sigma_1$ | 0.40 | (59; 40) | $\gamma_{IV2}$ | 0.125 | (59; 58; 67)* |
| $\eta_{PV1}$ | 1.25 | (40)* | $\sigma_{V1}$ | 0.40 | (59; 40)* | $\gamma_{A1}$ | 0.2 | (59; 40) |
| $\eta_{P2}$ | 1.25 | (40)* | $\sigma_2$ | 0.40 | (59; 40)* | $\gamma_{AV1}$ | 0.37 | (59; 40; 67)* |
| $\eta_{PV2}$ | 1.25 | (40)* | $\sigma_{V2}$ | 0.40 | (59; 40)* | $\gamma_{A2}$ | 0.2 | (59; 40)* |
| $\eta_{A1}$ | 0.75 | (18) | $\xi_1$ | 0.40 | (59; 40) | $\gamma_{AV2}$ | 0.33 | (59; 40; 67)* |
| $\eta_{AV1}$ | 0.75 | (18)* | $\xi_{V1}$ | 0.40 | (59; 40)* | $\gamma_{H1}$ | 0.0714 | (18) |
| $\eta_{A2}$ | 0.75 | (18)* | $\xi_2$ | 0.40 | (59; 40)* | $\gamma_{HV1}$ | 0.086 | (18; 67)* |
| $\eta_{AV2}$ | 0.75 | (18)* | $\xi_{V2}$ | 0.40 | (59; 40)* | $\gamma_{H2}$ | 0.0714 | (18)* |
| $\eta_{H1}$ | 0.25 | (40) | $r$ | 0.6 | (18) | $\gamma_{HV2}$ | 0.083 | (18; 67)* |
| $\eta_{HV2}$ | 0.25 | (40)* | $r_v$ | 0.1 | (24)* | $\delta_{I1}$ | 0.0008 | (40; 18) |
| $\eta_{H2}$ | 0.25 | (40)* | $q$ | 0.6 | (18)* | $\delta_{IV1}$ | 0 | (40; 18)* |
| $\eta_{HV2}$ | 0.25 | (40)* | $q_v$ | 0.6 | (18)* | $\delta_{I2}$ | 0.0015 | (40; 18)* |
| $\theta_v$ | 0.5 | (52; 67; 41) | $\phi_1$ | 0.02 | (38)* | $\delta_{IV2}$ | 0.0001 | (40; 18)* |
| $\theta_c$ | 0.75 | (52; 67; 41)* | $\phi_{V1}$ | 0.005 | (24; 14) | $\delta_{H1}$ | 0.0025 | (40) |
| $\psi_1$ | 0 | (40) | $\phi_2$ | 0.02 | (38) | $\delta_{HV1}$ | 0.0010 | (40)* |
| $\psi_{V1}$ | 0 | (40)* | $\phi_{V2}$ | 0.01 | (24; 14)* | $\delta_{H2}$ | 0.005 | (40; 45)* |
| $\psi_2$ | 0 | (40)* | $\gamma_{I1}$ | 0.1 | (59; 58) | $\delta_{HV2}$ | 0.0025 | (40; 45)* |
| $\psi_{V2}$ | 0 | (40)* | $\gamma_{IV1}$ | 0.13 | (59; 58; 67)* | $\omega_v$ | 0 | (40) |

\* indicates value was adapted from source based on current available data.

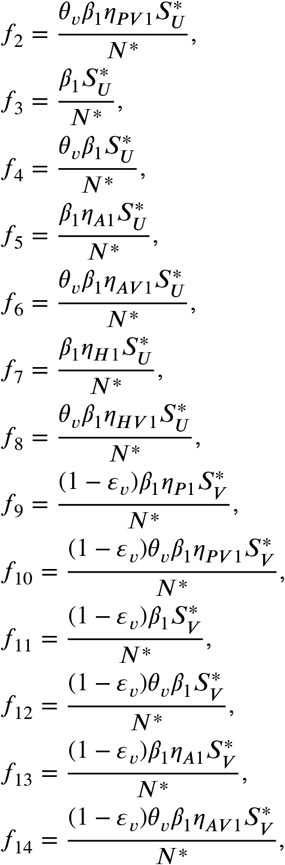

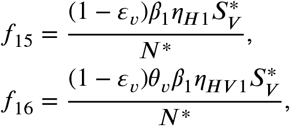

and

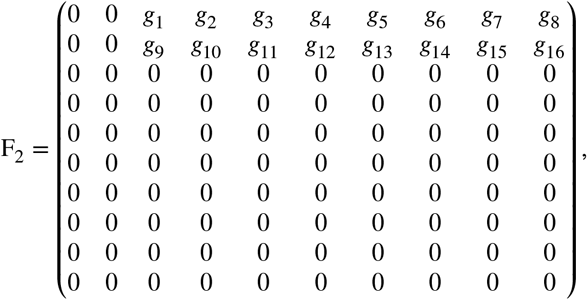

with entries,

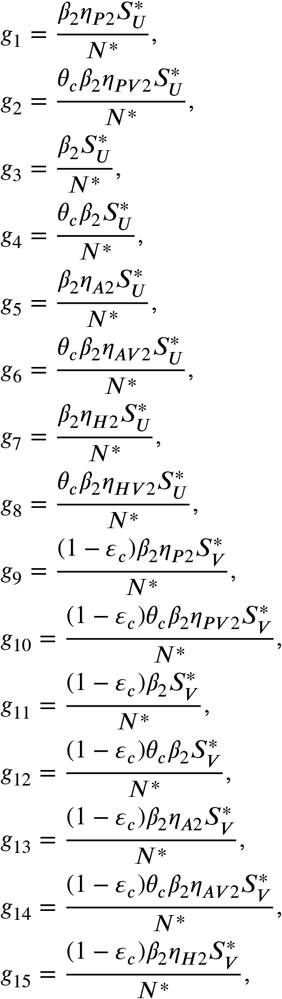

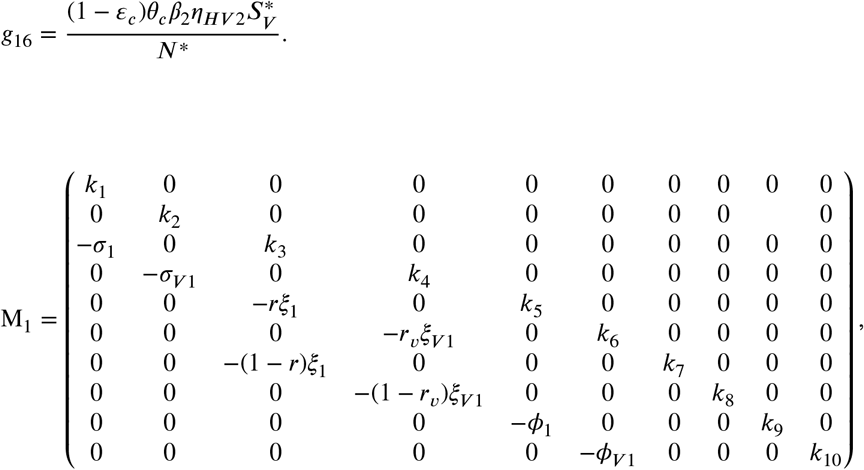

with entries,

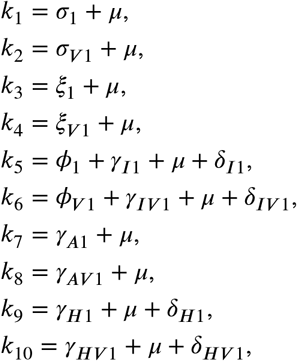

and

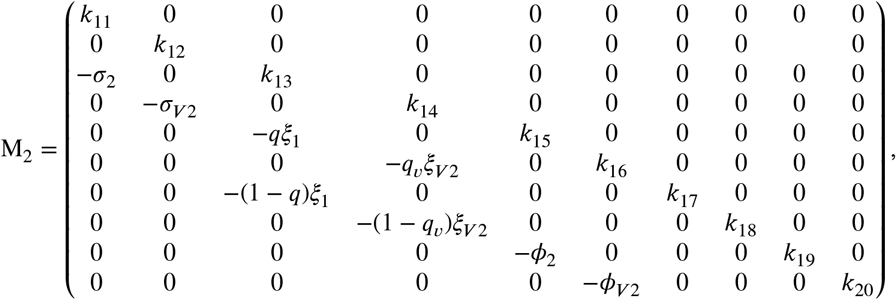

with entries,

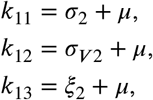

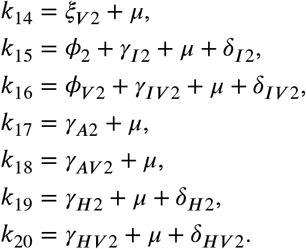

Using the approach in (39), the *vaccination reproduction numbers* of the wild-type (strain 1) and variant (strain 2) SARS-CoV-2 strains are found from

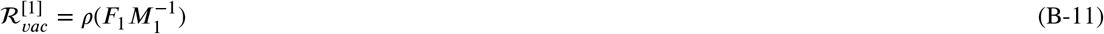

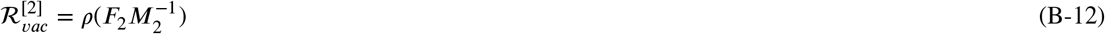

respectively, where *ρ* denotes the spectral radius. The expressions for 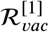 and 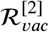 are given by:

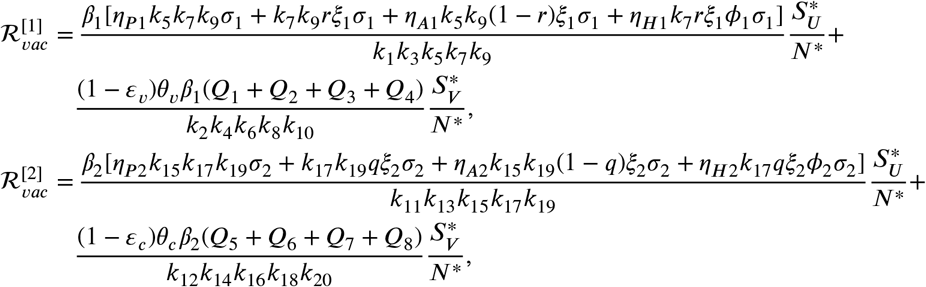

where,

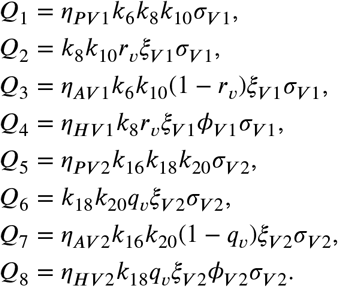

## References

[1] G. Abbott, (2020). Executive Order GA 14.

[2] G. Abbott, (2020). Executive Order GA 29.

[3] G. Abbott, (2020). Executive Order GA 34.

[4] Abu-Raddad L.J., A.A. Butt, (2021). Effectiveness of the BNT162b2 Covid-19 Vaccine against the B.1.1.7 and B.1.351 Variants. New England Journal of Medicine. doi: 10.1056/NEJMc2104974.

[5] Allen H., et al. (2021). Increased household transmission of COVID-19 cases associated with SARS-CoV-2 variant of concern B.1.617.2: a national case-control study. https://khub.net/documents/135939561/405676950/Increased+Household+Transmission+of+COVID-19+Cases+-+national+case+study.pdf/7f7764fb-ecb0-da31-77b3-b1a8ef7be9aa. Accessed August 3, 2021.

[6] Anderson R.M. (1992). The concept of herd immunity and the design of community-based immunization programmes. Vaccine 10(13), 928–935. doi: 10.1016/0264-410x(92)90327-g.

[7] Anderson, R.M., R.M. May (1985). Vaccination and herd immunity to infectious diseases, Nature 318(6044), 323–329. doi: 10.1038/318323a0

[8] Betti, M., N. Bragazzi, J. Heffernan, J. Kong, A. Raad (2021). Could a new COVID-19 mutant strain undermine vaccination efforts? A mathematical modelling approach for estimating the spread of B.1.1.7 using Ontario, Canada as a case study. Vaccines, 9, 592. doi: 10.3390/vac-cines9060592.

[9] Bloomberg (2021). More Than 2.42 Billion Shots Given: Covid-19 Tracker. https://www.bloomberg.com/graphics/covid-vaccine-tracker-global-distribution/. Accessed June 20, 2021.

[10] Bloomberg Covid-19 Vaccine Tracker Open Data (2021). Github repository. https://github.com/BloombergGraphics/covid-vaccine-tracker-data. Accessed June 17, 2021.

[11] Blower S.M., H. Dowlatabadi (1994). Sensitivity and Uncertainty Analysis of Complex Models of Disease Transmission: an HIV Model, as an Example. International Statistical Review. 62(2), 229–243. doi: 10.2307/1403510.

[12] Centers for Disease Control and Prevention (2021). The Advisory Committee on Immunization Practices’ Updated Interim Recommendation for Allocation of COVID-19 Vaccine – United States, December 2020. Morbidity and Mortality Weekly Report (MMWR). https://www.cdc.gov/mmwr/volumes/69/wr/mm695152e2.htm?s_cid=mm695152e2_w. Accessed June 17, 2021.

[13] Centers for Disease Control and Prevention (2021). COVID Data Tracker. https://covid.cdc.gov/covid-data-tracker/#datatracker-home. Accessed August 3, 2021.

[14] Centers for Disease Control and Prevention COVID-19 Vaccine Breakthrough Case Investigations Team (2021). COVID-19 vaccine breakthrough infections reported to CDC - United States, January 1-April 20, 2021. Morbidity and Mortality Weekly Report, 70(21).

[15] Centers for Disease Control and Prevention (2021). Emerging SARS-CoV-2 Variants. https://www.cdc.gov/coronavirus/2019-ncov/science/science-briefs/scientific-brief-emerging-variants.html?CDC_AA_refVal=https%3A%2F%2Fwww.cdc.gov%2Fcoronavirus%2F2019-ncov%2Fmore%2Fscience-and-research%2Fscientific-brief-emerging-variants.html. Accessed June 17, 2021.

[16] Centers for Disease Control and Prevention (2021). How COVID-19 Spreads. https://www.cdc.gov/coronavirus/2019-ncov/prevent-getting-sick/how-covid-spreads.html. Accessed July 27, 2021.

[17] Centers for Disease Control and Prevention (2021). Long-Term Effects. https://www.cdc.gov/coronavirus/2019-ncov/long-term-effects.html. Accessed June 17, 2021.

[18] Centers for Disease Control and Prevention (2021). Pandemic Planning Scenarios. https://www.cdc.gov/coronavirus/2019-ncov/hcp/planning-scenarios.html. Accessed June 17, 2021.

[19] Centers for Disease Control and Prevention (2021). SARS-CoV-2 Variant Classifications and Definitions. https://www.cdc.gov/coronavirus/2019-ncov/variants/variant-info.html?CDC_AA_refVal=https%3A%2F%2Fwww.cdc.gov%2Fcoronavirus%2F2019-ncov%2Fcases-updates%2Fvariant-surveillance%2Fvariant-info.html. Accessed August 3, 2021.

[20] Centers for Disease Control and Prevention (2021). Variant Proportions. https://covid.cdc.gov/covid-data-tracker/#variant-proportions. Accessed August 3, 2021.

[21] Center for Systems Science and Engineering at Johns Hopkins University. (2021). COVID-19. Github repository. https://github.com/CSSEGISandData/COVID-19. Accessed March 6, 2021.

[22] Colorado Governor Jared Polis (2020). Gov. Polis and State Public Health Officials Announce First Case of COVID Variant of COVID-19 in Colorado. https://www.colorado.gov/governor/news/3856-gov-polis-and-state-public-health-officials-announce-first-case-covid-variant-covid-19. Accessed June 17, 2021.

[23] Cuevas, E. (2020). An agent-based model to evaluate the covid-19 transmission risks in facilities. Computers in Biology and Medicine. 121, doi: 10.1016/103827.

[24] Dagan, N., et al. (2021). BNT162b2 mRNA covid-19 vaccine in a nationwide mass vaccination setting. New England Journal of Medicine. doi: 10.1056/NEJMoa2101765.

[25] Davies N.G., et al. (2020). Estimated transmissible and severity of novel SARS-CoV-2 Variant of Concern 202012/01 in England. medRxiv. https://cmmid.github.io/topics/covid19/reports/uk-novel-variant/2020_12_23_Transmissibility_and_severity_of_VOC_202012_01_in_England.pdf. Accessed March 6, 2021.

[26] Diekmann O., J.A.P. Heesterbeek, and J.A.J. Metz (1990) On the definition and the computation of the basic reproduction ratio R0 in models for infectious diseases in heterogenous populations. Journal of Mathematical Biology, 28, 365–382.

[27] Ducey, D. (2021). State of Arizona Executive Order 2021-06. March 25, 2021.

[28] Ducey, D. (2020). State of Arizona Executive Order 2020-18.

[29] Eikenberry S.E., et al. (2020). To mask or not to mask: Modeling the potential for face mask use by the general public to curtail the COVID-19 pandemic. Infectious Disease Modeling, 5, 293–308. DOI: 10.1016/j.idm.2020.04.001.

[30] Elbasha E.H., A.B. Gumel (2006). Theoretical Assessment of Public Health Impact of Imperfect Prophylactic HIV-1 Vaccines with Therapeutic Benefits. Bulletin of Mathematical Biology, 68, 577–614. doi: 0.1007/s11538-005-9057-5.

[31] Ferguson, N.M., et al. (2020). Impact of non-pharmaceutical interventions (NPIs) to reduce COVID-19 mortality and healthcare demand. London: Imperial College COVID-19 Response Team.

[32] Firth J.A., J. Hellewell, P. Klepac, S. Kissler (2020). Using a real-world network to model localized covid-19 control strategies. Nature Medicine, 26(10):1616–22. doi: 10.1038/s41591-020-1036-8.

[33] FOX 10 Phoenix (2020). LIST: Arizona cities with face mask requirements. https://www.fox10phoenix.com/news/list-arizona-cities-with-face-mask-requirements. Accessed June 17, 2021.

[34] Gonzalez-Parra, G., D. Martinez-Rodriguez & R.J. Villaneuva-Mico (2021). Math. Comput. Appl, 26, 25. doi: 10.3390/mca26020025.

[35] Gordon R., (2020). Gatherings and Face Mask Order. Michigan Department of Health and Human Services. Accessed June 17, 2021.

[36] Greaney, A.J., et al. (2021). Comprehensive mapping of mutations in the SARS-CoV-2 receptor-binding domain that affect recognition by polyclonal human plasma antibodies. Cell Host & Microbe. 29(3):463–476. doi: 10.1016/j.chom.2021.02.003.

[37] Greenhalgh, J. (2021). The Highly Contagious Delta Variant is On The Rise In The U.S. National Public Radio. https://www.npr.org/sections/coronavirus-live-updates/2021/06/08/1004597294/the-highly-contagious-delta-variant-of-covid-is-on-the-rise-in-the-u-s. Accessed June 17, 2021.

[38] Gumel, A.B., E.A. Iboi, c.n. Ngonghala, E.H. Elbasha (2021). A primer on using mathematics to understand COVID-19 dynamics: Modeling, analysis, and simulations. Infectious Disease Modelling, 6:1–21. doi: 10.1016/j.idm.2020.11.005.

[39] Gumel, A.B., B. Song (2008). Existence of Multiple-Stable Equilibria for a Multi-Drug-Resistant Model of Mycobaceria. Mathematical Biosciences and Engineering, 5(3): 437–455. doi: 0.3934/mbe.2008.5.437.

[40] Gumel, Abba B., Iboi, Enahoro, Ngonghala, Calistus and Ngwa, Gideon. Towards achieving a vaccine-derived herd immunity threshold for COVID-19 in the U.S. Frontiers in Public Health. doi: 10.3389/fpubh.2021.709369.

[41] Harris, R.J. et al.. (2021). Effect of vaccination on household transmission of SARS-CoV-2 in England. New England Journal of Medicine. doi: 10.1056/NEJMc2107717.

[42] Hellewell, J., et al. (2020). Feasibility of controlling COVID-19 outbreaks by isolation of cases and contacts. The Lancet Global Health 8(4):e488–e496. doi: 10.1016/S2214-109X(20)30074-7

[43] Hethcote, H.W. (2000). The mathematics of infectious diseases. SIAM Review, 42(4): 599–653.

[44] Hoertel, N., et al. (2020). A stoachastic agent-based model of the sars-cov-2 epidemic in France. Nature Medicine, 26:1417–1421.

[45] Horby, P., et al. (2021). NERVTAG note on B.1.1.7 severity for SAGE. New and Emerging Respiratory Virus threads Advisory Group.

[46] Iboi, E.A., C.N. Ngonghala, A.B. Gumel (2020). Will an imperfect vaccine curtail the COVID-19 pandemic in the US? Infectious Disease Modelling 5:510–524. doi: 10.1016/j.idm.2020.07.006.

[47] Impelli, M (2021). These cities are reviving mask mandates with new CDC guidance, delta surge. Newsweek. https://www.msn.com/en-us/news/us/these-cities-are-reviving-mask-mandates-with-new-cdc-guidance-delta-surge/ar-AAMF5N1. Accessed August 3, 2021.

[48] Institute for Health Metrics and Evaluation. (2020). Forecasting COVID-19 impact on hospital bed-days, ICU-days, ventilator-days, and deaths by US state in the next 4 months. MedRxiv.

[49] Klas, M.E., S. Contorno, (2020). Florida Gov. Ron DeSantis issues statewide stay-at-home order. Tampa Bay Times. https://www.tampabay.com/news/health/2020/04/01/florida-gov-ron-desantis-issues-statewide-stay-at-home-order/. Accessed June 17, 2021.

[50] Korber, B., et al. (2020) Tracking Changes in SARS-CoV-2 Spike: Evidence that D614G Increases Infectivity of the COVID-19 Virus. Cell, 182(4):812–827. doi: 10.1016/j.cell.2020.06.043.

[51] Kurcharski, A.J., et al. (2020). Early dynamics of transmission and control of covid-19: A mathematical modelling study. The Lancet Infectious Diseases, 20(5):553–558. doi: 10.1016/S1473-3099(20)30144-4.

[52] Levine-Tiefenbrun, M., et al. (2021). Initial report of decreased SARS-CoV-2 viral load after inoculation with the BNT162b2 vaccine. Nature Medicine, 7: 790–792. doi: 10.1038/s41591-021-01316-7.

[53] Lovelace Jr., B. & N. Rattner (2021). CDC says 7-day average of daily U.S. covid cases surpassed peak seen last summer. CNBC. https://www.cnbc.com/2021/08/02/cdc-says-7-day-average-of-daily-us-covid-cases-surpassed-peak-seen-last-summer.html. Accessed August 3, 2021.

[54] Mayo Clinic (2021). COVID-19 vaccines: Get the facts. https://www.mayoclinic.org/coronavirus-vaccine/art-20484859. Accessed June 17, 2021.

[55] McKay, M.D., R.J. Beckan, W.J. Conover (1979). A Comparison of Three Methods for Selecting Values of Input Variables in the Analysis of Output from a Computer Code. Technometrics, 21(2): 239–245. doi: 10.2307/1271432.

[56] McLeod R.G., J.F. Brewster, A.B. Gumel, D.A. Slonowsky (2006). Sensitivity and Uncertainty Analyses for a SARS Model with Time-Varying Inputs and Outputs. Mathematical Biosciences and Engineering, 3(3): 27–544. doi: 10.3934/mbe.2006.3.527.

[57] Minnesota Department of Health (2021). MDH lab testing confirms nation’s first known COVID-19 case associated with Brazil P.1 variant. https://www.health.state.mn.us/news/pressrel/2021/covid012521.html. Accessed June 17, 2021.

[58] Ngonghala, C.N., et al. (2020). Mathematical assessment of the impact of non-pharmaceutical interventions on curtailing the 2019 novel Coronavirus. Mathematical Biosciences, 325. doi: 10.1016/j.mbs.2020.108364

[59] Ngongahala, C.N., E. Iboi, A.B. Gumel (2020). Could masks curtail the post-lockdown resurgence of covid-19 in the US? Mathematical Biosciences, 329. doi: 10.1016/j.mbs.2020.108452

[60] National Institute of Allergies and Infectious Diseases (2020). COVID-19 MERS, & SARS. National Institute of Health. https://www.niaid.nih.gov/diseases-conditions/covid-19. Accessed June 17, 2021.

[61] Polletta, M., A. Oxford (2020). Arizona governor issues ‘stay-at-home’ order; will take effect close of business Tuesday. azcentral. https://www.azcentral.com/story/news/local/arizona-health/2020/03/30/arizona-coronavirus-stay-home-order-issued-gov-doug-ducey/5088109002/. Accessed June 17, 2021.

[62] Skelly, D.T., et al. (2021). Vaccine-inuced immunity provides more robust heterotypic immunity than natural infection to emerging SARS-CoV-2 variants of concern. Research Square. https://www.researchsquare.com/article/rs-226857/v1. doi: 10.21203/rs.3.rs-226857/v1

[63] South Carolina Department of Health and Environmental Control (2021). South Carolina Public Health Officials Detect Nation’s First Known Cases of the COVID-19 Variant Originally Detected in South Africa. https://scdhec.gov/index.php/news-releases/south-carolina-public-health-officials-detect-nations-first-known-cases-covid-19. Accessed June 17, 2021

[64] Srivastava, A. and G. Chowell (2020). Understanding spatial heterogeneity of COVID-19 pandemic using shape analysis of growth rate curves. MedRxiv. doi: 10.1101/2020.05.25.20112433.

[65] Sullivan, B. (2021). U.S. COVID deaths are rising again. Experts call it a ‘pandemic of the unvaccinated’. National Public Radio. https://www.npr.org/2021/07/16/1017002907/u-s-covid-deaths-are-rising-again-experts-call-it-a-pandemic-of-the-unvaccinated. Accessed August 3, 2021.

[66] Tariq, A., et al. (2020). Real-time monitoring the transmission potential of COVID-19 in Singapore, March 2020. BMC Medicine, 18, 1–14.

[67] Thompson, M.G., et al. (2021). Prevention and Attenuation of covid-19 with the BNT162b2 and mRNA-1273 vaccines. New England Jounral of Medicine, 385:320–9. doi: 10.1056/NEJMoa2107058.

[68] Thurner, S., P. Klimek, R. Hanel (2020). A network-based explanation of why most covid-19 infection curves are linear. Proceedings of the National Academy of Sciences, 117(37): 22684–22689. doi: 10.1073/pnas.2010398117.

[69] United States Census Bureau, (2020). National Demographic Analysis Tables: 2020. https://www.census.gov/data/tables/2020/demo/popest/2020-demographic-analysis-tables.html. Accessed June 17, 2021.

[70] United States Food and Drug Administration (2021). Coronavirus (COVID-19) Update: FDA Issues Policies to Guide Medical Product Developers Addressing Virus Variants. https://www.fda.gov/news-events/press-announcements/coronavirus-covid-19-update-fda-issues-policies-guide-medical-product-developers-addressing-virus. Accessed June 17, 2021.

[71] United States Food and Drug Administration (2020). FDA Briefing Document Moderna COVID-19 Vaccine. https://www.fda.gov/media/144434/download. Accessed June 17, 2021.

[72] United States Food and Drug Administration (2020). FDA Briefing Document Pfizer-BioNTech COVID-19 Vaccine. https://www.fda.gov/media/144245/download. Accessed June 17, 2021.

[73] United States Food and Drug Administration (2021). FDA Issues Emergency Use Authorization for Third COVID-19 Vaccine. https://www.fda.gov/news-events/press-announcements/fda-issues-emergency-use-authorization-third-covid-19-vaccine. Accessed June 17, 2021.

[74] United States Food and Drug Administration (2020). FDA Takes Key Action in Fight Against COVID-19 By Issuing Emergency Use Authorization for First COVID-19 Vaccine. https://www.fda.gov/news-events/press-announcements/fda-takes-key-action-fight-against-covid-19-issuing-emergency-use-authorization-first-covid-19. Accessed June 17, 2021.

[75] United States Food and Drug Administration. Pfizer-BioNTech COVID-19 Vaccine. https://www.fda.gov/emergency-preparedness-and-response/coronavirus-disease-2019-covid-19/pfizer-biontech-covid-19-vaccine. Accessed June 17, 2021.

[76] van den Driessche P., J. Watmough (2002). Reproduction numbers and sub-threshold endemic equilibria for compartmental models of disease transmission, Mathematical Biosciences, 180:29–48. doi: 10.1016/S0025-5564(02)00108-6.

[77] Volz, E., et al. (2021). Transmission of SARS-CoV-2 Lineage B.1.1.7 in England: Insights from linking epidemiological and genetic data. MedRxiv. doi: 10.1101/2020.12.30.20249034.

[78] WESH 2, (2020). Florida governor extends order suspending COVID-19 related enforcement fines. https://www.wesh.com/article/florida-covid-19-mask-fines-order-extended/34778032. Accessed June 17, 2021

[79] Whitmer, G. (2020). Executive Order No. 2020-42. State of Michigan Office of the Governor.

[80] Whitmer, G. (2020). Executive Order No. 2020-60. State of Michigan Office of the Governor.

[81] Whitmer, G. (2020). Executive Order No. 2020-110. State of Michigan Office of the Governor. https://www.michigan.gov/coronavirus/0,9753,7-406-98178_98455-550215--,00.html.

[82] World Health Organization (2020). WHO Diretor-General’s opening remakrs at the mission briefing on COVID-19. https://www.who.int/director-general/speeches/detail/who-director-general-s-opening-remarks-at-the-mission-briefing-on-covid-19.

[83] Wu, K. et al.. (2021). mRNA-1273 vaccine induces neutralizing antibodies against spike mutants from global SARS-CoV-2 variants. biorxiv. doi: 10.1101/2021.01.25.427948.

[84] Xie, X., et al. (2021). Neutralization of SARS-CoV-2 spike 69/70 deletion, E484K, and N501Y variants by BNT162b2 vaccine-elicited sera. bioRxiv. doi: 10.1038/s41591-021-01270-4.

[85] Xue, L., et al. (2020). A data-driven network model for the emerging covid-19 epidemics in Wuhan, Toronto, and Italy. Mathematical Biosciences, 326. doi: 10.1016/j.mbs.2020.108391.

[86] Zhu, N. et al.. (2020). A novel coronavirus from patients with pneumonia in China, 2019. New England Journal of Medicine 382(8): 727–733. doi: 10.1056/NEJMoa2001017.

